# Shared and ethnic background site-specific dietary patterns in the Hispanic Community Health Study/Study of Latinos (HCHS/SOL)

**DOI:** 10.1101/2022.06.30.22277013

**Authors:** Roberta De Vito, Briana Stephenson, Daniela Sotres-Alvarez, Anna-Maria Siega-Riz, Josiemer Mattei, Maria Parpinel, Brandilyn A. Peters, Sierra A. Bainter, Martha L. Daviglus, Linda Van Horn, Valeria Edefonti

## Abstract

Dietary patterns (DPs) synthesize multiple related dietary components in one or more combined variables. A drawback of DPs is their limited reproducibility across subpopulations, especially adopting *a posteriori* DPs, derived using standard multivariate methods [e.g., factor analysis (FA)]. Standard approaches assessing reproducibility of FA-based DPs mostly rely on correlation coefficients/agreement measures between pairs of factors and do not consider any statistical model. Multi-study factor analysis builds upon standard FA model to identify DPs shared across all subpopulations and those specific to some subpopulations. Pattern reproducibility is investigated from a different perspective: a shared DP identified within multi-study factor analysis is “reproducible” since it is common to all subpopulations. Bayesian multi-study factor analysis (BMSFA) has been developed to improve DP retention and identification, two critical issues as the number of subpopulations analyzed increases.

Using baseline (2008-2011) 24-hour dietary recalls from the Hispanic Community Health Study/Study of Latinos (n=16,415), we applied the BMSFA on 42 common nutrients to identify shared and subpopulation-specific DPs where subpopulations were defined as the cross-classification of ethnic background and study site (EBS).

Overall, 4 shared DPs were identified: *Plant-based foods, Processed foods, Dairy products*, and *Seafood*. At the subpopulation level, we identified 12 EBS-specific DPs, one for each EBS category, primarily representing variants of foods from animal sources. Different nuances were expressed by subsets of fairly similar EBS-specific DPs, including an *<u>Animal vs. vegetable source</u>*, an *<u>Animal source only</u>*, and a *<u>Poultry vs. dairy products</u>* overarching DPs. Shared DPs from BMSFA were similar to their counterparts from standard FA and frequentist multi-study factor analysis; EBS-specific DPs from BMSFA were better characterized than those from frequentist multi-study factor analysis.

In conclusion, the BMSFA successfully captured sources of both dietary homogeneity and heterogeneity in a large well-characterized study of US Hispanics/Latino adults by ethnic background and site.

## Introduction

*A posteriori* dietary patterns (DPs) (1), defined by using multivariate statistical methods [i.e., principal component analysis (PCA), factor analysis (FA), and cluster analysis (2)], are advantageous in naturally reflecting actual study- or population-specific context (e.g., geography/climate, socioeconomic status, food supply, ethnic background, religion) (3), but their specificity limits generalizability, compared to the *a priori* (i.e., comparing subjects’ diet against evidence-based benchmark diets) options (4). The absence of a standardized approach to analysis has further hindered discovery of genuinely reproducible *a posteriori* DPs across subpopulations within the same study or other studies from the same or similar populations (i.e., *cross-study* reproducibility (5)). The two issues have traditionally limited comparisons among sets of *a posteriori* DPs, thereby preventing firm conclusions about their health benefits (or risks) (6, 7).

To our knowledge, only one paper (8) has explored reproducibility of *a posteriori* DPs in subpopulations belonging to the same study. This study used stratified PCAs by US region, sex, and race to choose the optimal number of components to retain for a final analysis based on the overall sample. In a multi-cultural population, a full exploration of regional and ethnic diversity is important, but it requires efficient statistical methods to manage and compare the different level-specific solutions (9). Yet, different ethnic backgrounds pose additional research challenges, including identification of a proper food grouping scheme and coping with possible masked acculturation effects on dietary habits (10).

A few pioneering (11, 12) and more recent (13-19) papers have explored *cross-study reproducibility* of PCA-based or FA-based DPs across different studies. One of two major statistical approaches is applied on a common set of variables across studies (3): 1. a stratified approach, i.e., each study expresses its own set of DPs and a reproducibility analysis is carried out using congruence/correlation coefficients or agreement measures (13, 14, 18); or 2. a pooled approach, i.e., studies are merged in a single dataset and forced to share common DPs (15, 16).

Multi-study factor analysis (20) (MSFA), recently applied in nutritional epidemiology (17), identifies shared DPs, common to all studies, as well as additional study-specific DPs for individual studies. Shared DPs identified within MSFA are “reproducible” across studies since they are common to all studies. Estimation of MSFA model parameters using Bayesian techniques [Bayesian multi-study factor analysis (BMSFA)] reportedly improve factor identification and choice of the number of factors to retain (21). These are two issues of great importance as the number of studies/subpopulations analyzed increases.

The Hispanic Community Health Study/Study of Latinos (HCHS/SOL) (22) is an ongoing US multi-site community-based cohort studying health and risk factors of cardiovascular and pulmonary outcomes of Hispanic/Latino adults, and differences across several ethnic backgrounds. This cohort provides the unique opportunity to define both shared and subpopulation-specific DPs by the cross-section of ethnic background and site (EBS), in support of future culturally tailored interventions. Two papers within HCHS/SOL have identified shared and subpopulation-specific *a posteriori* DPs using approaches conceptually similar to the MSFA (23, 24). Stephenson et al. (24) used robust profile clustering to cluster participants and food items (from the food propensity questionnaire) based on consumption behaviors shared amongst all participants and those specific to 9 EBS subpopulations (24). Compared to robust profile clustering, MSFA derives DPs analogous to those from FA or PCA, providing a more straightforward interpretation for nutrition researchers. In addition, robust profile clustering has limited functionality in handling continuous data like nutrient intakes. Maldonado et al. (23) conducted stratified principal FAs by 6 ethnic backgrounds on a common set of food groups from 24-hour recalls. Overarching DPs were defined based on food groups that shared high loadings in multiple ethnic backgrounds, but DP similarity was not statistically evaluated. Even though investigator-consensus was used to reduce inherent subjectivity, overall factor selection is cumbersome to manage when several stratified FAs are performed, influencing the number and composition of the identified overarching DPs. In addition, associations of background-specific DPs with health outcomes are difficult to interpret. Our MSFA derives reproducible DPs directly, improving factor retention through BMSFA, and easing interpretability of associations with health outcomes of interest.

This paper aims to explore the use of BMSFA to jointly identify shared and EBS-specific DPs from a common set of nutrients derived from 24-hour recalls to address the following research questions: 1. Are there consistent and empirically estimable DPs shared across the HCHS/SOL? 2. Are there one or more additional DPs for some EBS-specific categories? 3. Are the identified shared and EBS-specific DPs interpretable in terms of food groups, and socio-demographic and lifestyle factors?

## Materials and methods

### Study design

The HCHS/SOL enrolled 16,415 adults aged 18–74 years residing in four US field sites (Bronx, Chicago, Miami, San Diego) from six Hispanic/Latino ethnic backgrounds (Cuban, Dominican Republic, Mexican, Puerto Rican, Central and South American) at baseline (2008-2011). A stratified 2-stage area probability design was adopted. Sampling design and cohort selection have been described previously (25). Participants provided written informed consent, and the Institutional Review Boards from each field center and the University of North Carolina at Chapel Hill (Coordinating Center) approved the study.

### Protocol and Measurements

The baseline visit included questionnaires administered in Spanish or English based on the participant’s language preference, anthropometry, and two 24-hour dietary recalls, among many other procedures. The first recall was administered in person and the 2^nd^ one via telephone, preferably ≤30 days from baseline in the participant’s preferred language by trained interviewers using the Nutrition Data System for Research software (version 11) (26). Virtually all participants (99%) provided at least one recall.

### Selection of participants

From the original cohort (n=16,415), participants were excluded who self-identified as belonging to other/multi/mixed (n=938) ethnic backgrounds, had recall data deemed unreliable by the interviewer (n=65), or provided extreme (i.e., <1st or >99th sex-specific percentiles) estimates of energy intake (n=171). After these exclusions, we removed 184 participants belonging to EBS categories with <200 participants – “fair” sample size to carry out FA (27) (i.e., South American - Bronx) – and 36 with missing Hispanic/Latino background. Overall, 15,021 participants were used for analysis from 12 EBS categories (**Supplemental Figure 1**).

### Specification of variables and data preprocessing

From the Nutrition Data System for Research list of 139 available nutrients (26), we selected 42 that best represent the overall diet for Hispanics/Latinos; we expanded the fat profile to capture cardiovascular disease-related dietary habits (28, 29). Only nutrient intake from foods, not from supplements, was considered. We derived final intake from one or the mean of two available reliable recalls. Intakes were all log-transformed (base e) to improve the normality of the shared and EBS-specific factors, and of the EBS-specific errors (details in **Supplemental Methods)**.

### Statistical analysis

#### Factorability of the correlation matrices

We used Bartlett’s test of sphericity, overall and individual measures of sampling adequacy (27), to assess whether the 12 EBS-specific correlation matrices and the overall correlation matrix of the log-transformed nutrient intakes were factorable.

#### Identification of nutrient-based dietary patterns

We carried out BMSFA (21) on the EBS-specific log-transformed data correlation matrices to estimate unobservable shared (K) and EBS-specific (J_s_) factors, known as DPs. Compared with the frequentist approach (20) [here named frequentist multi-study factor analysis (FMSFA)], BMSFA offers two advantages: 1. a better-defined loading structure via the prior distribution that makes loadings extreme, and 2. a practical and useful approach (eigenvalue decomposition and 5% variance explained cut-off) to choose the number of shared and EBS-specific factors. After estimating the factor loadings, we applied the varimax rotation to the shared factor-loading matrix to obtain a better-defined loading structure. We named ‘dominant nutrients’ (27) those showing shared rotated (or EBS-specific unrotated) factor loadings **≥**|0.60| (**≥**|0.30|). Factor scores estimate the degree to which each participant’s diet is summarized by each identified DP. We calculated factor scores in BMSFA by following FA’s standard Thurstone approach (30, 31) (correlation with Bartlett method ≥0.90 for the shared factors).

We evaluated the internal reliability of DPs in two ways. First, we assessed internal consistency with standardized Cronbach’s alpha and *alpha-when-item-deleted* coefficients for those nutrients that loaded >|0.40| on the shared (>|0.20| on the EBS-specific) factors (27). Second, we assessed internal reproducibility by comparing BMSFA-based DPs with those from principal component FA (PCFA) on the overall sample (16) and FMSFA (17, 20). Throughout the paper, we referred to the congruence coefficient (CC) and its cut-offs proposed in (14) for DP comparison: “fair similarity” corresponds to 0.85≥CC≤0.94 and “equivalence” to CC>0.95 (**Supplemental Methods** for additional details).

#### Interpreting dietary patterns in terms of food groups, socio-demographic and lifestyle characteristics

We separately classified participants based on factor scores’ quintiles (shared factors) or tertiles (EBS-specific factors) to validate DPs against selected 24-hour-recall-based food groups, socio-demographic and lifestyle factors. For the food-group-based validation, we estimated adjusted mean intakes of food groups (servings/day), stratified by quantile-based categories of DPs, using a linear regression model adjusted for age, sex, body mass index (BMI), EBS category (for shared factors only), and total energy intake. We expressed the deviation of the food group intake relative to its overall (i.e., based on the total final sample size) or EBS-specific mean as: *100%×[adjusted mean within quintile-based category/overall HCHS/SOL mean]* for the shared factors and *100% ×[adjusted mean within tertile-based category/EBS-specific mean]* for the EBS-specific factors. Therefore, if the consumption of a food group exceeded 100%, individuals from that factor score quantile category were characterized by high consumption of that food group, compared with the HCHS/SOL overall (shared factor) or EBS-specific (EBS-specific factor) mean, and vice versa when the relative intake is below 100%.

For the validation of DPs against selected socio-demographic and lifestyle characteristics, we used the Pearson Chi-square test of independence (categorical characteristics) or the ANOVA (continuous characteristics); adjustment for multiple comparisons was carried out with the False Discovery Rate Method (32). To provide a comparison between the identified DPs and the Alternative Healthy Eating Index (AHEI-2010) – a measure of overall diet quality previously related to major chronic disease risk and mortality, with strong associations with cardiometabolic disease (11 components, score range: 0-110, lowest to highest quality]) (33) – we additionally estimated the adjusted mean AHEI-2010 score within quantile-based categories of factor scores using a linear regression model adjusted for age, sex, BMI, EBS combination (for shared factors only), and total energy intake. We calculated the p-values for linear trend based on the same models.

Except for the implementation of BMSFA, all statistical analyses accounted for HCHS/SOL complex survey design, including survey weights, stratification and clustering (34). All statistical tests were two-sided. Calculations were carried out using the open-source statistical computing environment R (35), with its libraries “statmod” (36), “psych” (37), “nFactors” (38), “ggplot2” (39), “survey” (40), and “MSFA” (41).

## Results

### Population characteristics

Socio-demographic and lifestyle characteristics by EBS categories were included in **Table 1**. The largest and smallest EBS categories were participants of Mexican background from San Diego (n=3775) and Bronx (n=205), respectively. Across all EBS categories, most individuals were between the ages of 18-44 years and reported an income of less than $30,000; in most EBS categories, the majority of individuals were first-generation immigrants and non-consumers of supplements (all p-values<0.001). Differences between EBS categories were observed for sex, years living in the United States, age of immigration, marital status, employment status, education, physical activity, BMI, and energy intake. The highest percentage of individuals born in mainland US was identified for the Cuban background – Miami category, which also had the highest percentage of not meeting the 2008 Physical Activity Guidelines for Americans and the highest mean energy intake. Individuals of Mexican background from the Bronx had the highest percentage of being 18– 44 years old and married, receiving an income <$30,000, and following the 2008 Physical Activity Guidelines for Americans. They also showed almost the lowest mean energy intake. The Mexican background – Chicago individuals had the highest AHEI-2010 mean value, whereas individuals of Puerto Rican backgrounds from Chicago and the Bronx had the lowest AHEI-2010 scores, and the lowest mean age at arrival at mainland US. The Mexican background – San Diego category had the highest percentage of individuals in the >$30,000 income category. The South American background - Miami category had the highest percentage of highly educated individuals and individuals who used dietary supplements.

**Table 1.**
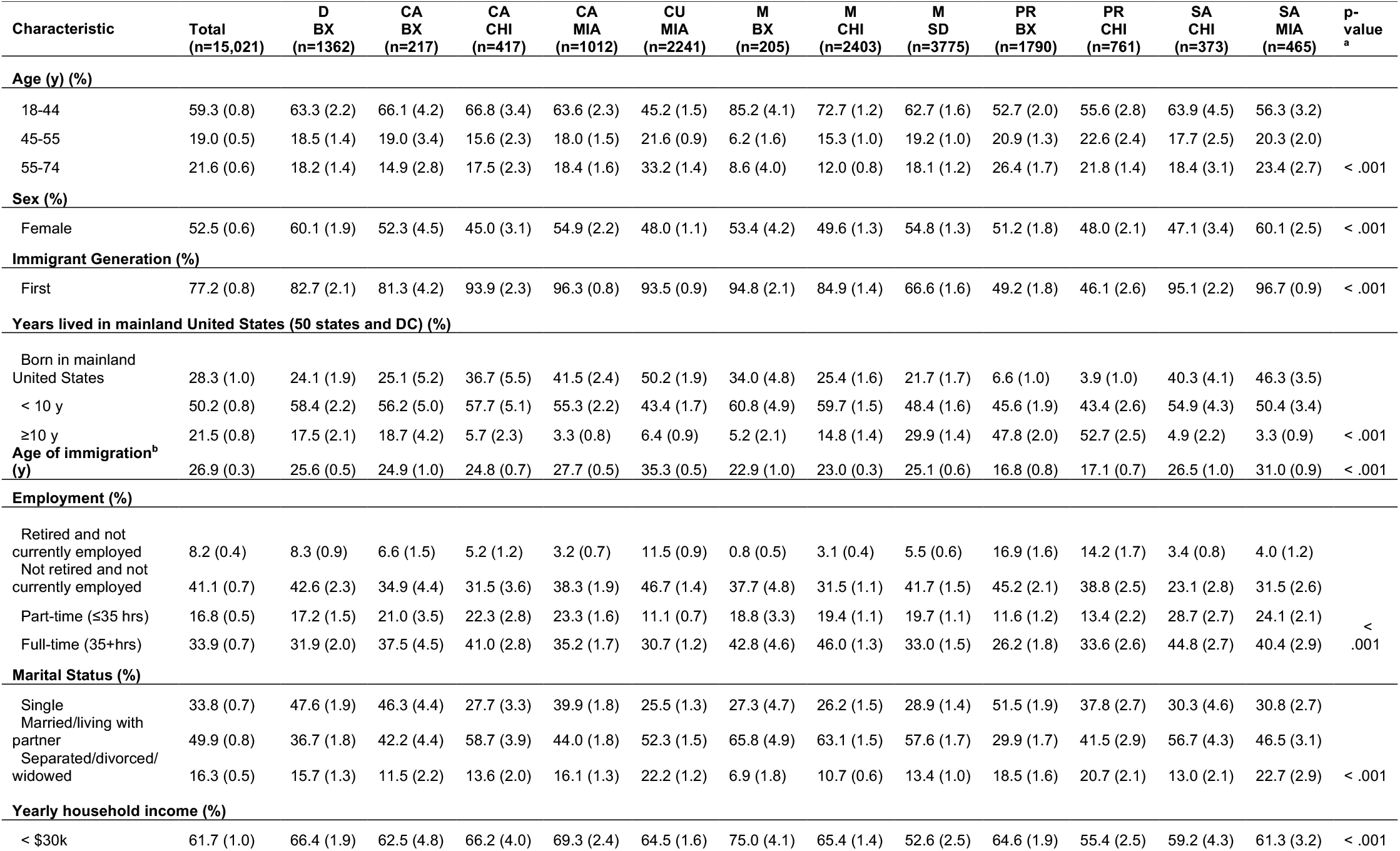
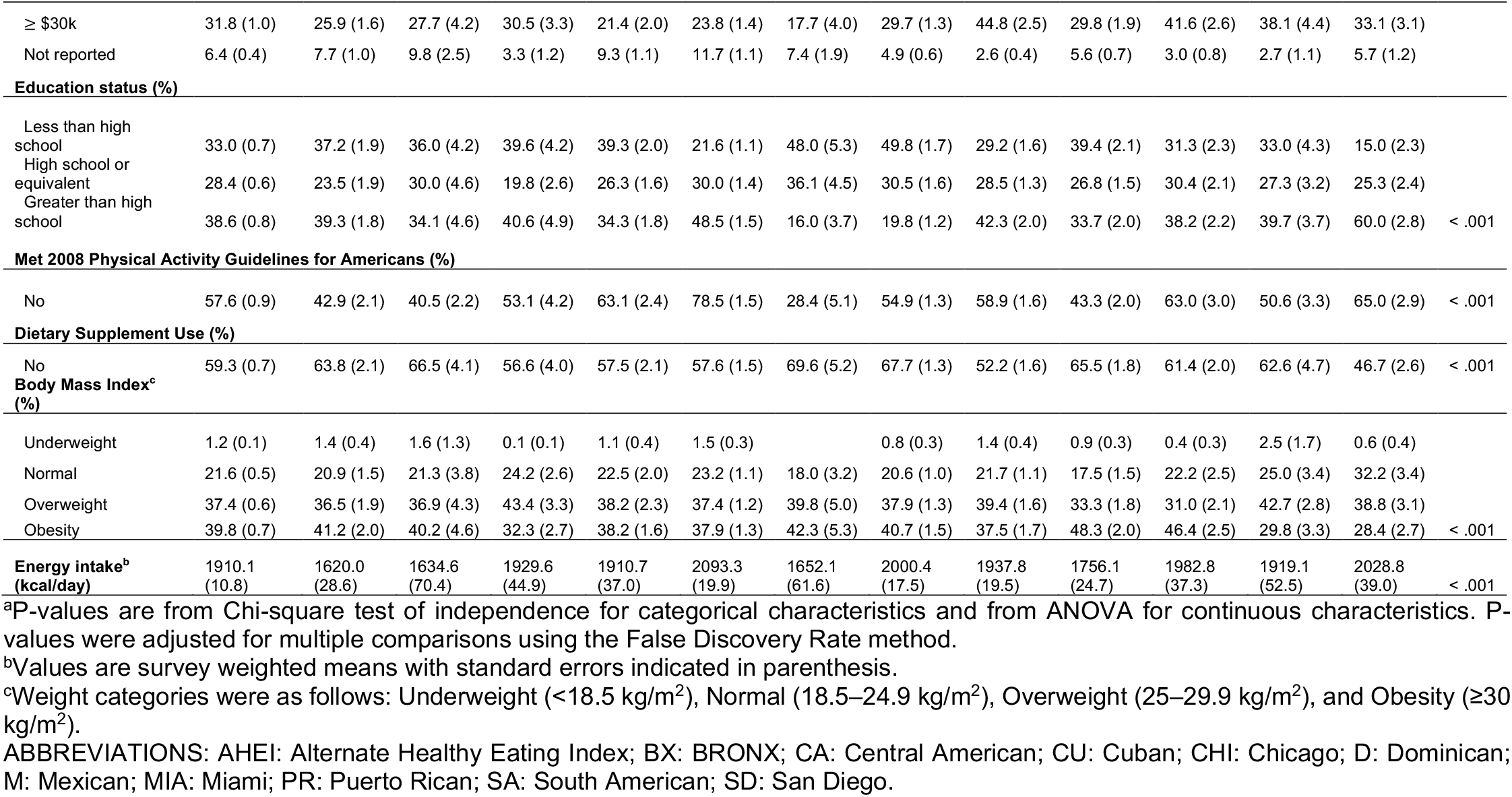
Baseline socio-demographic and lifestyle characteristics (weighted percentages and standard error in parenthesis) by ethnic background and study site (EBS). Hispanic Community Health Study/Study of Latinos, 2008-2011.

### Identification of nutrient-based dietary patterns

Factorability of correlation matrices was confirmed for each of the 12 EBS-specific and overall correlation matrices **(Supplemental Results** and **Supplemental Table 1**). The BMSFA estimated 4 shared DPs, common to all EBS categories (explaining 62.5% of the total variance, **Supplemental Table 2**) and one EBS-specific DP for each of the 12 categories (explaining a variance ranging from 10.7% for Central American background – Miami to 14.4% for Puerto Rican background – Chicago, **Supplemental Table 3**). A heatmap illustrates the factor loadings for shared and EBS-specific DPs (**Figure 1**).

**Figure 1.**
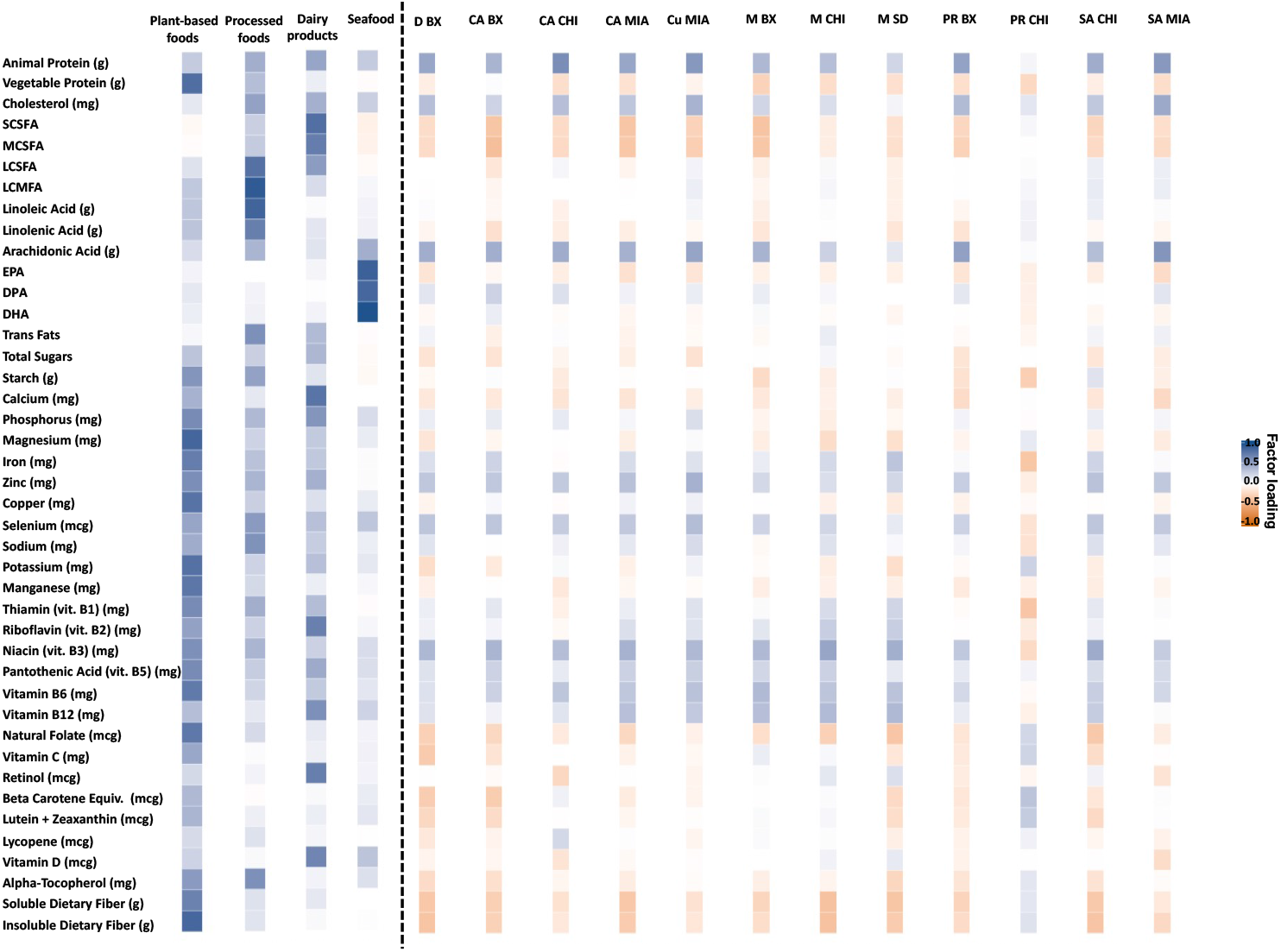
Heatmap of the estimated factor-loading matrix for the shared and ethnic-background-site specific dietary patterns identified with the BMFA. Dashed line indicates the division between the shared and ethnic-background-site-specific dietary patterns. Hispanic Community Health Study/Study of Latinos, 2008-2011. ABBREVIATIONS: BMSFA: Bayesian multi-study factor analysis; BX: BRONX; CA: Central American; Cu: Cuban; CHI: Chicago; D: Dominican; DHA: docosahexaenoic acid; DPA: docosapentaenoic acid; EPA: eicosapentaenoic acid; M: Mexican; MCSFA: medium-chain saturated fatty acids; LCMFA: long-chain monounsaturated fatty acids; LCSFA: long-chain saturated fatty acids; MIA: Miami; PR: Puerto Rican; SA: South American; SCSFA: short-chain saturated fatty acids; SD: San Diego.

### Shared dietary patterns

Factor 1, namely *Plant-based foods*, showed high (i.e., dark blue in **Figure 1**, values in **Supplemental Table 2**) factor loadings on vegetable protein, phosphorus, magnesium, iron, zinc, copper, potassium, manganese, thiamin, niacin, pantothenic acid, vitamin B6, natural folate, soluble and insoluble dietary fiber. Factor 2, namely *Processed foods*, showed high loadings on long-chain saturated fatty acids, long-chain monounsaturated fatty acids, linoleic acid, linolenic acid, total trans fatty acids, and natural alpha-tocopherol. Factor 3, namely *Dairy products*, showed high loadings on short-and medium-chain saturated fatty acids, calcium, riboflavin, vitamin B12, retinol, and vitamin D. Factor 4, namely *Seafood*, showed high loadings on eicosapentaenoic acid (EPA), docosapentaenoic acid (DPA), and docosahexaenoic acid (DHA).

### Ethnic background-site specific (EBS) dietary patterns

Ten of the identified EBS-specific DPs represented variants of an animal profile, as they were characterized by 2 or 3 nutrients among animal protein, arachidonic acid, and niacin (dark blue color in **Figure 1**). Based on CC (**Table 2**) and visual inspection of the factor loadings (**Supplementary Table 3**), additional profile similarities were related to common ethnic background or site and were expressed through the following three overarching EBS-specific DPs (i.e., DPs shared among some EBS-specific categories):

**Table 2.**
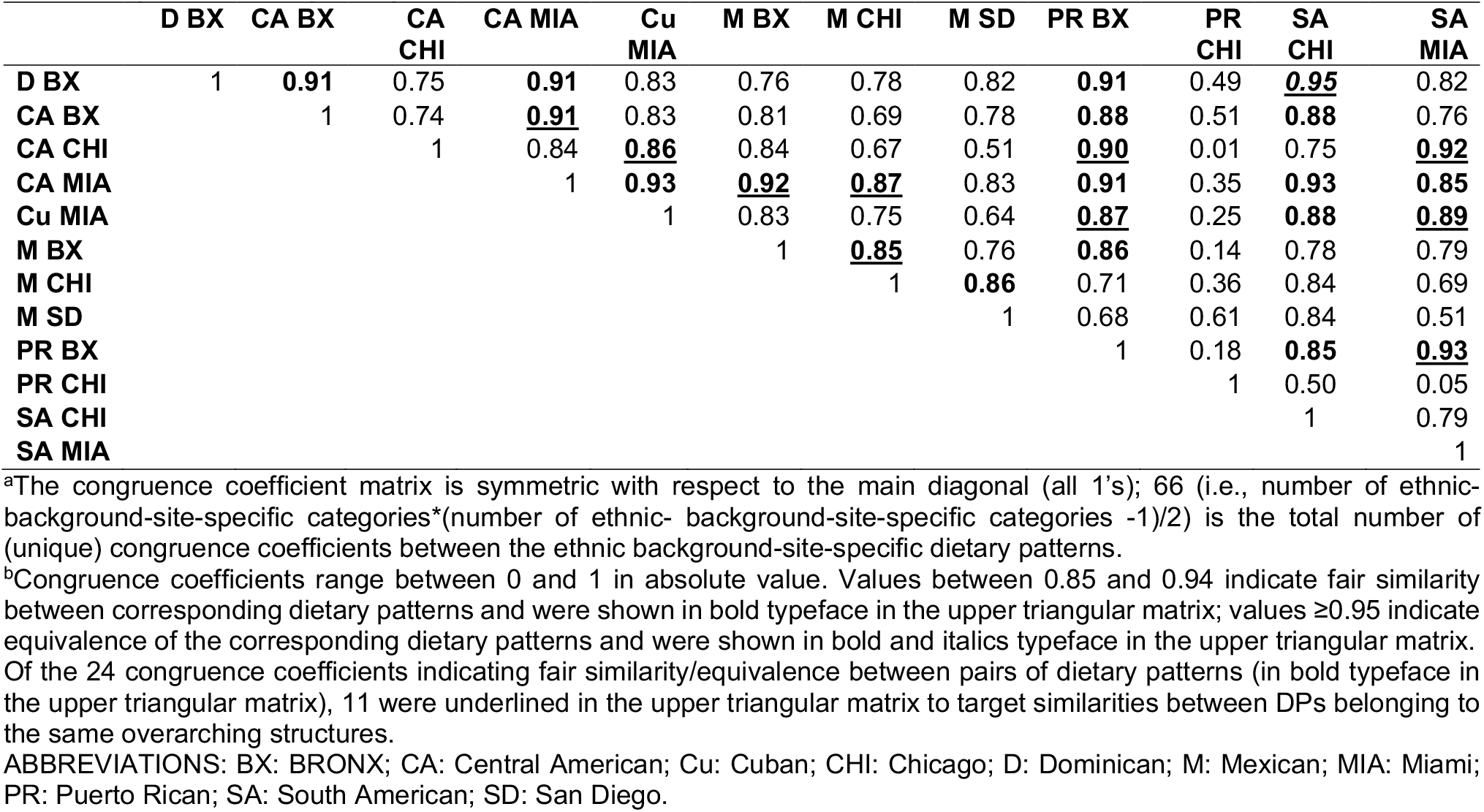
Factor congruence coefficients^a,b^ between pairs of ethnic-background-site-specific dietary patterns. Hispanic Community Health Study/Study of Latinos, 2008-2011.

|  | D BX | CA BX | CA CHI | CA MIA | Cu MIA | M BX | M CHI | M SD | PR BX | PR CHI | SA CHI | SA MIA |
| --- | --- | --- | --- | --- | --- | --- | --- | --- | --- | --- | --- | --- |
| D BX | 1 | **0.91** | 0.75 | **0.91** | 0.83 | 0.76 | 0.78 | 0.82 | **0.91** | 0.49 | **<u>0.95</u>** | 0.82 |
| CA BX |  | 1 | 0.74 | **<u>0.91</u>** | 0.83 | 0.81 | 0.69 | 0.78 | **0.88** | 0.51 | **0.88** | 0.76 |
| CA CHI |  |  | 1 | 0.84 | **<u>0.86</u>** | 0.84 | 0.67 | 0.51 | **<u>0.90</u>** | 0.01 | 0.75 | **<u>0.92</u>** |
| CA MIA |  |  |  | 1 | **0.93** | **<u>0.92</u>** | **<u>0.87</u>** | 0.83 | **0.91** | 0.35 | **0.93** | **0.85** |
| Cu MIA |  |  |  |  | 1 | 0.83 | 0.75 | 0.64 | **<u>0.87</u>** | 0.25 | **0.88** | **<u>0.89</u>** |
| M BX |  |  |  |  |  | 1 | **<u>0.85</u>** | 0.76 | **0.86** | 0.14 | 0.78 | 0.79 |
| M CHI |  |  |  |  |  |  | 1 | **0.86** | 0.71 | 0.36 | 0.84 | 0.69 |
| M SD |  |  |  |  |  |  |  | 1 | 0.68 | 0.61 | 0.84 | 0.51 |
| PR BX |  |  |  |  |  |  |  |  | 1 | 0.18 | **0.85** | **<u>0.93</u>** |
| PR CHI |  |  |  |  |  |  |  |  |  | 1 | 0.50 | 0.05 |
| SA CHI |  |  |  |  |  |  |  |  |  |  | 1 | 0.79 |
| SA MIA |  |  |  |  |  |  |  |  |  |  |  | 1 |

<sup>a</sup>The congruence coefficient matrix is symmetric with respect to the main diagonal (all 1's); 66 (i.e., number of ethnic-background-site-specific categories\*(number of ethnic-background-site-specific categories -1)/2) is the total number of (unique) congruence coefficients between the ethnic background-site-specific dietary patterns.
<sup>b</sup>Congruence coefficients range between 0 and 1 in absolute value. Values between 0.85 and 0.94 indicate fair similarity between corresponding dietary patterns and were shown in bold typeface in the upper triangular matrix; values $\geq 0.95$ indicate equivalence of the corresponding dietary patterns and were shown in bold and italics typeface in the upper triangular matrix. Of the 24 congruence coefficients indicating fair similarity/equivalence between pairs of dietary patterns (in bold typeface in the upper triangular matrix), 11 were underlined in the upper triangular matrix to target similarities between DPs belonging to the same overarching structures.
ABBREVIATIONS: BX: BRONX; CA: Central American; Cu: Cuban; CHI: Chicago; D: Dominican; M: Mexican; MIA: Miami; PR: Puerto Rican; SA: South American; SD: San Diego.

#### (1) Animal vs. vegetable source

animal protein/arachidonic acid *vs*. soluble and insoluble dietary fiber (CC reached 0.95, meaning equivalence, between DPs of Dominican background – Bronx and South American background – Chicago);

#### (2) Animal source only

animal protein/arachidonic acid/cholesterol with the same sign *vs*. no other dominant nutrients (0.85≥CC≤0.94, meaning fair similarity, for DPs of the following pairs: Puerto Rican background – Bronx, Central American background – Chicago, Cuban background – Miami, and South American background – Miami);

#### (3) Poultry vs. dairy products

less clearly identified and characterized by combinations of animal protein/arachidonic acid/niacin/vitamin B6/vitamin B12 *vs*. small- and medium-chain saturated fatty acids/soluble and insoluble fiber (0.85≥CC≤0.94, meaning fair similarity for 4 out of 6 possible pairs of EBS-specific DPs among the following: Central American background – Bronx, Central American background – Miami, Mexican background – Bronx, Mexican background – Chicago).

In contrast, one EBS-specific category, Puerto Rican background – Chicago, was characterized by a strikingly different DP, high on beta-carotene (dark blue color) and low on starch, iron, and thiamin (orange color) (**Figure 1**), which showed no fair similarity with any other EBS-specific DP. In addition, the Mexican background – San Diego DP loaded high on iron, niacin, vitamin B6, vitamin B12, as opposed to natural folate, soluble and insoluble fiber (**Figure 1**), showing fair similarity with the Mexican background – Chicago DP only (**Table 2**).

Finally, of the 24 CCs (36%) indicating fair similarity/equivalence between pairs of EBS-specific DPs in the upper triangular matrix (**Table 2**), 11 (underlined in the table) targeted similarities between DPs belonging to the same overarching structure and 13 targeted similarities between DPs belonging to different overarching structures, thus suggesting similarities also across overarching DPs.

### Internal reproducibility and consistency of the identified patterns

Nutrient communalities and internal consistency of DPs, measured by Cronbach’s alpha, were satisfactory, further supporting our selection of nutrients (**Supplementary Table 4** and **Supplemental Results**). The internal reproducibility of BMSFA-based DPs – as based on the comparison with FMSFA and PCFA – was reassuring (**Figure 2, Supplemental Results** and **Supplemental Tables 5-8**). Indeed, BMSFA estimated the same number of DPs and a similar total variance explained under FMSFA and PCFA, although DP order differed in PCFA, with a more prominent role of the *Processed foods* DP (**Supplemental Table 5**). The BMSFA-based shared DPs were equivalent to their counterparts from FMSFA (all CCs ≥0.97) and PCFA (all CCs ≥0.95) (**Figure 2**, which shows a heatmap of CCs between the shared DPs identified with the BMSFA and those from PCFA and FMSFA). Visual inspection of the factor-loading matrix for the shared DPs (**Supplemental Table 5**) suggested that BMSFA was more effective in: 1. shrinking/increasing moderately low/high loadings in absolute value towards 0 or 1; 2. forcing potentially dominant nutrients to load on one DP instead of two.

**Figure 2.**
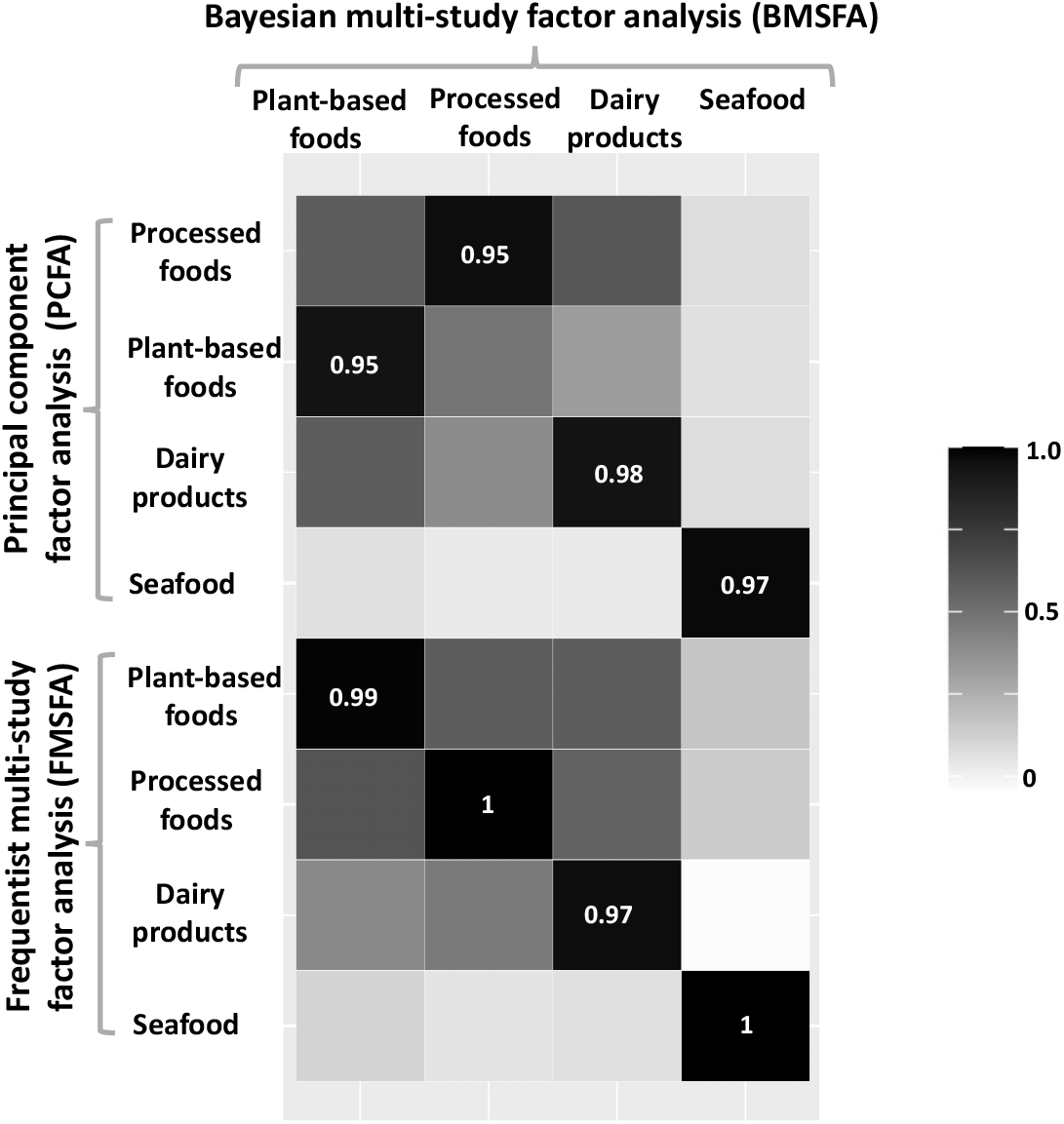
Heatmap of the factor congruence coefficients^a,b^ between the shared dietary patterns identified with the BMSFA and FMSFA and the overall-sample dietary patterns from PCFA. Hispanic Community Health Study/Study of Latinos, 2008-2011. ^a^Congruence coefficients range between 0 and 1 (in absolute value), with values between 0.85 and 0.94 indicating fairly similarity and values ≥0.95 indicating equivalence of the corresponding dietary patterns. ^b^Dietary patterns were ordered in terms of proportion of total variance explained in each solution (see **Supplemental Table 5**).

While percentages of explained variances were similar to their BMSFA-specific counterparts, FMSFA-specific DPs generally opposed animal and vegetable sources of foods (animal protein/cholesterol/arachidonic acid *vs*. vegetable protein/folate/soluble and insoluble fiber in ≥8 DPs, **Supplemental Table 6**) and therefore showed fewer nuances than the corresponding BMSFA-based ones (fair similarity/equivalence for FMSFA: 92% *vs*. 36% for BMSFA, **Supplemental Table 7** *vs*. **Table 2**). This was confirmed in the one-to-one comparison between FMSFA-and BMSFA-based versions of the same EBS-specific DP, with 4 DPs only being fairly similar under the two approaches (**Supplemental Table 8)**.

### Top consumers of shared dietary patterns by ethnic background-site category

Ethnic background-site categories were well represented in the top quintile category of each shared DP, with a prevalence (i.e., weighted percentage) around the expected 20% (i.e., 18-22%) for most EBS categories (**Supplemental Table 9**). Major deviations from the 18-22% interval were observed for the *Seafood* DP, with percentages as low as 13.1% (Mexican background – Bronx) and as high as 24.3% (South American background – Chicago).

### Food groups associated with the identified dietary patterns

#### Shared dietary patterns

**Table 3** shows the deviation (%) of the adjusted food group mean for individuals in the top quintile of each shared DP, relative to the overall adjusted mean. Individuals in the top quintile category of the *Plant-based foods* DP were characterized by high (i.e., >140%) intakes of fruit (citrus and other) and vegetables (dark green, orange, tomatoes, beans, and others), whole grain, nuts and seeds, and alcohol, compared to the overall HCHS/SOL mean. Individuals in the top quintile category of *Processed foods* DP were characterized by high intakes of starchy vegetables, refined grain, red meat, deli meat, eggs, nuts and seeds, cheese, milk-based desserts, sugar sweetened beverages, alcohol, snacks, and added fats (regular and reduced). Individuals in the top quintile category of the *Dairy products* DP showed high consumption of deli meats, milk, cheese, yogurt, and milk-based desserts; they also showed a low (i.e., <60%) consumption of nuts and seeds. Finally, individuals in the top quintile category of the *Seafood* DP presented an extremely high fish intake, and high consumption of dark green vegetables, poultry, and alcohol.

**Table 3.**
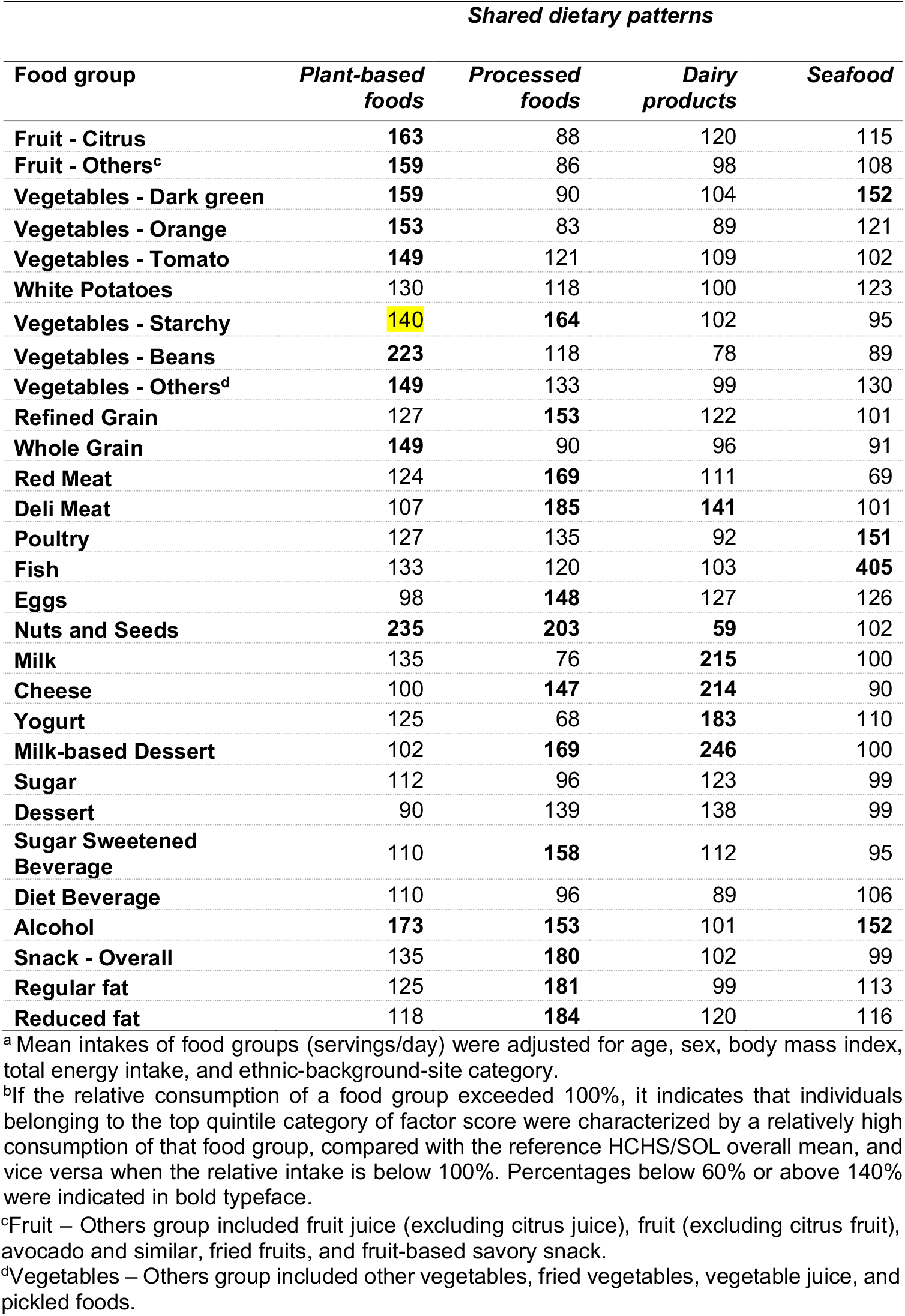
**Deviation (%) of the adjusted food group mean for individuals in the top quintile of each shared dietary pattern, relative to the overall adjusted mean. Hispanic Community Health Study/Study of Latinos, 2008-2011. ^a,b^**

| *Shared dietary patterns* |  |  |  |  |
| --- | --- | --- | --- | --- |
| **Food group** | ***Plant-based foods*** | ***Processed foods*** | ***Dairy products*** | ***Seafood*** |
| **Fruit - Citrus** | **163** | 88 | 120 | 115 |
| **Fruit - Others<sup>c</sup>** | **159** | 86 | 98 | 108 |
| **Vegetables - Dark green** | **159** | 90 | 104 | **152** |
| **Vegetables - Orange** | **153** | 83 | 89 | 121 |
| **Vegetables - Tomato** | **149** | 121 | 109 | 102 |
| **White Potatoes** | 130 | 118 | 100 | 123 |
| **Vegetables - Starchy** | **140** | **164** | 102 | 95 |
| **Vegetables - Beans** | **223** | 118 | 78 | 89 |
| **Vegetables - Others<sup>d</sup>** | **149** | 133 | 99 | 130 |
| **Refined Grain** | 127 | **153** | 122 | 101 |
| **Whole Grain** | **149** | 90 | 96 | 91 |
| **Red Meat** | 124 | **169** | 111 | 69 |
| **Deli Meat** | 107 | **185** | **141** | 101 |
| **Poultry** | 127 | 135 | 92 | **151** |
| **Fish** | 133 | 120 | 103 | **405** |
| **Eggs** | 98 | **148** | 127 | 126 |
| **Nuts and Seeds** | **235** | **203** | **59** | 102 |
| **Milk** | 135 | 76 | **215** | 100 |
| **Cheese** | 100 | **147** | **214** | 90 |
| **Yogurt** | 125 | 68 | **183** | 110 |
| **Milk-based Dessert** | 102 | **169** | **246** | 100 |
| **Sugar** | 112 | 96 | 123 | 99 |
| **Dessert** | 90 | 139 | 138 | 99 |
| **Sugar Sweetened Beverage** | 110 | **158** | 112 | 95 |
| **Diet Beverage** | 110 | 96 | 89 | 106 |
| **Alcohol** | **173** | **153** | 101 | **152** |
| **Snack - Overall** | 135 | **180** | 102 | 99 |
| **Regular fat** | 125 | **181** | 99 | 113 |
| **Reduced fat** | 118 | **184** | 120 | 116 |

<sup>a</sup> Mean intakes of food groups (servings/day) were adjusted for age, sex, body mass index, total energy intake, and ethnic-background-site category.
<sup>b</sup> If the relative consumption of a food group exceeded 100%, it indicates that individuals belonging to the top quintile category of factor score were characterized by a relatively high consumption of that food group, compared with the reference HCHS/SOL overall mean, and vice versa when the relative intake is below 100%. Percentages below 60% or above 140% were indicated in bold typeface.
<sup>c</sup>Fruit – Others group included fruit juice (excluding citrus juice), fruit (excluding citrus fruit), avocado and similar, fried fruits, and fruit-based savory snack.
<sup>d</sup>Vegetables – Others group included other vegetables, fried vegetables, vegetable juice, and pickled foods.

#### Ethnic background-site-specific (EBS) dietary patterns

Compared with each EBS-specific mean, participants in the top tertile category of most EBS-specific DPs showed a higher-than-mean consumption of poultry and alcohol *vs*. a lower-than-mean consumption of nuts and seeds, thus confirming the general animal source of most EBS-specific DPs. Red meat was also highly consumed in the top tertile category of half of the EBS-specific DPs. Overarching DPs additionally showed:

#### 1. Animal vs. vegetable source

higher-than-mean consumption of poultry *vs*. lower-than-mean consumption of non-citrus fruit (Dominican background – Bronx) or beans (South American background – Chicago);

#### 2. Animal source only

higher-than-mean consumption of red meat (except for South American background – Miami, but 132% was close to 140%) *vs*. no clear lower-than-mean consumption of other food groups;

#### 3. Poultry vs. dairy products

higher-than-mean consumption of poultry *vs*. lower-than-mean consumption of one or more among cheese, yogurt, and dairy dessert products, but still less clearly identified; lower-than-mean consumption of dairy products was common to individuals of Central American background from all sites and those of Cuban background from Miami, who belonged to the *<u>Animal source only</u>* overarching DP.

Individuals of Puerto Rican background – Chicago category in the top tertile showed a higher-than-mean consumption of yogurt, milk-based dessert products, (non-citrus) fruit, dark green and orange vegetables; this was different from any other EBS-specific DP, including that of their Bronx counterparts of Puerto Rican background. Finally, individuals of Mexican background from Chicago and San Diego showed a similar pattern of consumption, including red meat, poultry, and alcohol, but fewer nuts and seeds, and beans. However, no similarities were found with the few Mexican background individuals from Bronx, whose top-consumers showed high consumption of citrus fruit and sugar-sweetened beverages (**Table 4**).

**Table 4.** **Deviation (%) of the adjusted food group mean for individuals in the top tertile of each ethnic-background-site-specific dietary pattern, relative to the ethnic-background-site-specific adjusted mean. Hispanic Community Health Study/Study of Latinos, 2008-2011.^a,b^**

| *Ethnic background-site-specific (EBS) dietary patterns* |  |  |  |  |  |  |  |  |  |  |  |  |
| --- | --- | --- | --- | --- | --- | --- | --- | --- | --- | --- | --- | --- |
| **Food group** | **D BX** | **CA BX** | **CA CHI** | **CA MIA** | **Cu MIA** | **M BX** | **M CHI** | **M SD** | **PR BX** | **PR CHI** | **SA CHI** | **SA MIA** |
| **Fruit - Citrus** | 82 | 79 | 108 | 96 | 93 | **175** | 94 | 64 | 66 | 135 | 76 | 134 |
| **Fruit - Other<sup>c</sup>** | **56** | 60 | 74 | 83 | 84 | 117 | 77 | 71 | 96 | **151** | 77 | 89 |
| **Vegetables - Dark green** | 66 | **42** | 129 | 72 | 91 | 73 | 91 | 60 | 71 | **205** | 65 | 125 |
| **Vegetables - Orange** | 81 | 80 | 116 | 105 | 93 | 103 | 105 | 69 | 81 | **147** | 83 | 127 |
| **Vegetables - Tomato** | 83 | 80 | 106 | 105 | 110 | 124 | 94 | 89 | 79 | 110 | 91 | 97 |
| **White Potatoes** | 88 | 113 | **157** | 119 | 123 | 128 | 82 | 91 | 105 | 101 | 91 | 110 |
| **Vegetables - Starchy** | 86 | 106 | 80 | 82 | 90 | 72 | 114 | 91 | 90 | 126 | 94 | 104 |
| **Vegetables - Beans** | 100 | 107 | 82 | 62 | 81 | 64 | **43** | **44** | 85 | 98 | **56** | **59** |
| **Vegetables – Other<sup>d</sup>** | 81 | 78 | 112 | 111 | 106 | 123 | 101 | 82 | 88 | 118 | 81 | 101 |
| **Refined Grain** | 121 | 99 | 93 | 106 | 102 | 94 | 119 | 109 | 93 | 65 | 116 | 98 |
| **Whole Grain** | 79 | 84 | 108 | 93 | 94 | **44** | 73 | 86 | 81 | 115 | 81 | 100 |
| **Red Meat** | **157** | 134 | **150** | **149** | **160** | 138 | **141** | 138 | **152** | 94 | 137 | 132 |
| **Deli - Meat** | 124 | 67 | 92 | 114 | 113 | 93 | 114 | 100 | 113 | 74 | **145** | 119 |
| **Poultry** | **156** | **178** | **190** | **144** | **142** | **142** | **150** | 129 | **153** | 105 | **143** | **167** |
| **Fish** | 80 | 105 | 106 | 64 | 77 | 137 | 86 | 75 | **55** | 72 | 77 | 63 |
| **Eggs** | 111 | 124 | 93 | 119 | 126 | 94 | 98 | 82 | 125 | 104 | 95 | **150** |
| **Nuts and Seeds** | **18** | **16** | **20** | **53** | **47** | **5** | **55** | 62 | 65 | 105 | **28** | **36** |
| **Milk** | 76 | 78 | 66 | 95 | 93 | 96 | 90 | 103 | 81 | 105 | 91 | 79 |
| Food group | D BX | CA BX | CA CHI | CA MIA | Cu MIA | M BX | M CHI | M SD | PR BX | PR CHI | SA CHI | SA MIA |
| **Cheese** | 84 | **58** | **57** | 62 | 76 | 70 | 80 | 69 | 87 | 84 | **47** | 93 |
| **Yogurt** | 67 | **0** | 90 | 139 | 80 | 109 | 69 | 93 | **46** | **151** | **27** | 85 |
| **Milk-based Dessert** | 65 | **47** | **59** | **43** | **57** | 80 | **177** | 67 | 100 | **150** | 75 | 80 |
| **Sugar** | 86 | 87 | 87 | 74 | 82 | 78 | 83 | 87 | 93 | 108 | 82 | 98 |
| **Dessert** | 88 | 67 | 100 | **42** | 68 | 83 | 135 | 91 | 67 | 94 | 64 | 97 |
| **Sugar Sweetened Beverage** | 120 | 113 | 113 | 111 | 97 | **153** | 127 | 128 | 106 | 76 | 114 | 104 |
| **Diet Beverage** | 103 | 102 | 95 | 88 | 96 | 100 | 107 | 82 | 90 | 100 | 100 | 100 |
| **Alcohol** | 116 | 90 | **228** | **196** | **143** | **282** | **173** | **150** | **160** | 90 | **158** | 106 |
| **Snack - Overall** | 63 | 71 | 65 | 65 | 65 | **58** | 87 | 76 | 64 | 88 | 71 | 76 |
| **Regular fat** | 103 | 82 | 106 | 94 | 105 | 89 | 102 | 76 | 91 | 97 | 110 | 101 |
| **Reduced fat** | 100 | 116 | 87 | 105 | 101 | 83 | 129 | 104 | 94 | 102 | 111 | 103 |

<sup>a</sup> Mean intakes of selected food groups (servings/day) were adjusted for age, sex, body mass index, and total energy intake.
<sup>b</sup> If the relative consumption of a food group exceeded 100%, it indicates that individuals belonging to top tertile category of factor score were characterized by a relatively high consumption of that food group, compared with the ethnic background site specific overall mean, and vice versa when the relative intake is below 100%. Percentages below 60% or above 140% were indicated in bold typeface.
<sup>c</sup> Fruit – Others group included fruit juice (excluding citrus juice), fruit (excluding citrus fruit), avocado and similar, fried fruits, and fruit-based savory snack.
<sup>d</sup> Vegetables – Others group included other vegetables, fried vegetables, vegetable juice, and pickled foods.

### Alternative Healthy Eating Index associated with the Identified patterns

#### Shared dietary patterns

Higher quintile-based categories of *Plant-based foods* and *Seafood* DPs were consistently and significantly associated with higher mean AHEI-2010 scores (p for trend<0.001, 5-point increment from lowest to highest category for both DPs), suggesting a higher quality of top-consumers’ overall diet. *Dairy products* and *Processed foods* DPs showed the opposite trend (p for trend<0.001): lower mean AHEI-2010 scores (i.e., lower quality) were consistently reported for increasing quintile-based categories of these DPs, with an overall ∼3- and ∼5-point decrease, respectively (**Figure 3**).

**Figure 3.**
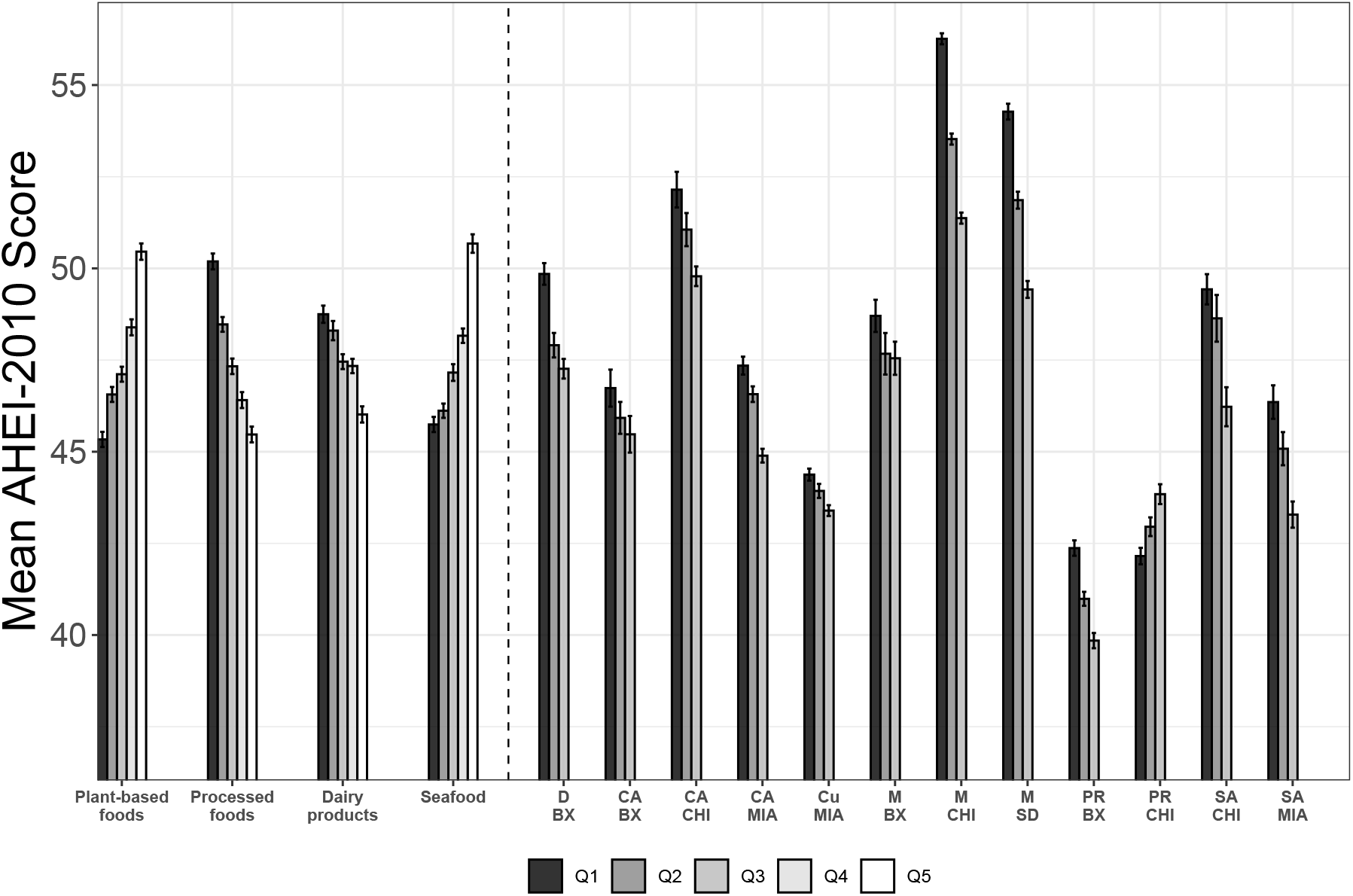
Mean AHEI-2010 scores (standard errors) by quintiles of shared dietary patterns and by tertiles of ethnic-background-site-specific dietary patterns^a,b,c^. Hispanic Community Health Study/Study of Latinos, 2008-2011. ^a^ Weighted mean AHEI-2010 scores were adjusted for age, sex, body mass index, and total energy intake; models for the shared dietary patterns were further adjusted for ethnic-background-site combinations and for the other shared dietary patterns (quintiles). Asterisks indicate significant p for trend: ***<0.001, **<0.01, and *<0.05, respectively. ^b^ The AHEI-2010 measures overall diet quality in terms of adherence to the Dietary Guidelines for Americans 2010; compared to the healthy Eating Index 2010, it also incorporates additional components that focus on foods and nutrients to predict the risk of chronic disease. The total AHEI-2010 score ranges from 0 to 110, with higher scores indicating a healthier diet. ^c^ The same color scale was adopted for the first three quintile-based categories and for the three tertile-based categories. ABBREVIATIONS: AHEI: Alternative Healthy Eating Index; BX: BRONX; CA: Central American; Cu: Cuban; CHI: Chicago; D: Dominican; M: Mexican; MIA: Miami; PR: Puerto Rican; SA: South American; SD: San Diego.

#### Ethnic background-site-specific (EBS) dietary patterns

Lower mean AHEI-2010 scores were consistently observed for increasing tertile-based categories of 11 EBS-specific combinations (all p for trend <0.05, except for Central American background – Bronx category: p=0.12). In contrast, mean AHEI-2010 scores globally increased by ∼2 points for increased consumption in participants of Puerto Rican background from Chicago. For the same tertile-based category, mean AHEI-2010 scores markedly differed across EBS-specific DPs, with Puerto Rican background participants from Bronx and Chicago showing the lowest mean scores [39.85 (SE: 0.20) and 42.15 (SE: 0.22), respectively] and Mexican background participants from Chicago and San Diego showing the highest mean scores [56.26 (SE: 0.15) and 54.27 (SE: 0.22), respectively] (**Figure 3**).

### Socio-demographic and lifestyle factors associated with the Identified patterns

#### Shared dietary patterns

While top-consumers of all shared DPs were more likely to be younger and males, percentages of the youngest or male participants were even more extreme in top-consumers of the *Processed foods* DP. Notably, top-consumers of the *Plant-based foods* and *Seafood* DPs were less likely to have spent more than 10 years in the US, whereas those of the *Dairy products* and *Processed foods* DPs were more likely to have spent more than 10 years in the US. Top-consumers of the *Processed foods* DP showed a lower weighted mean age at immigration. While most of the overall population did not meet the 2008 Physical Activity Guidelines for Americans, top-consumers of the *Plant-based foods* and *Processed foods* DPs were more likely to be active, than the lowest consumers. Use of supplements was more likely in the top-consumers of the *Plant-based foods* and *Dairy products* DPs and less likely in top-consumers of the *Processed foods* DP (**Table 5**, with most adjusted p-values<0.05).

**Table 5.**
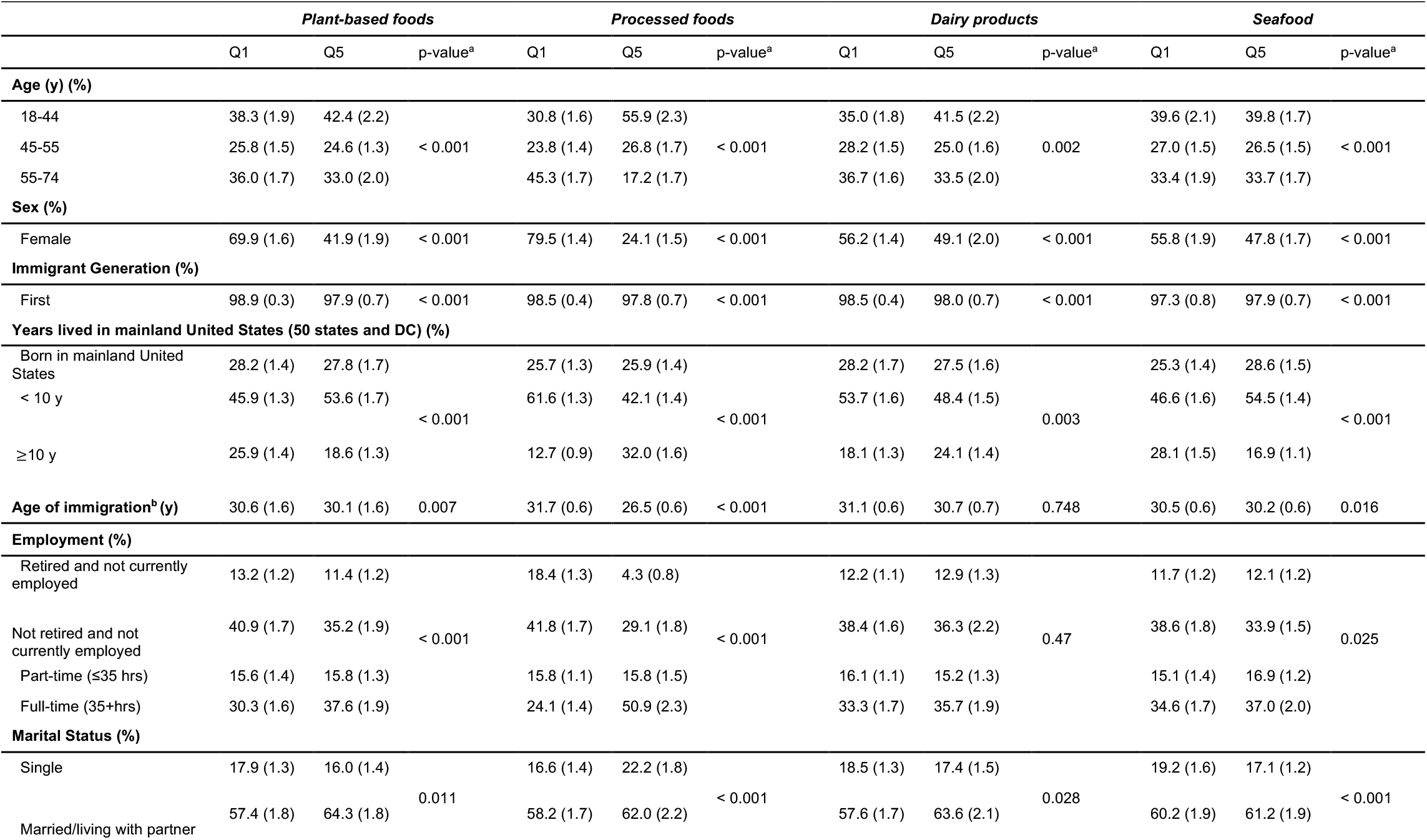
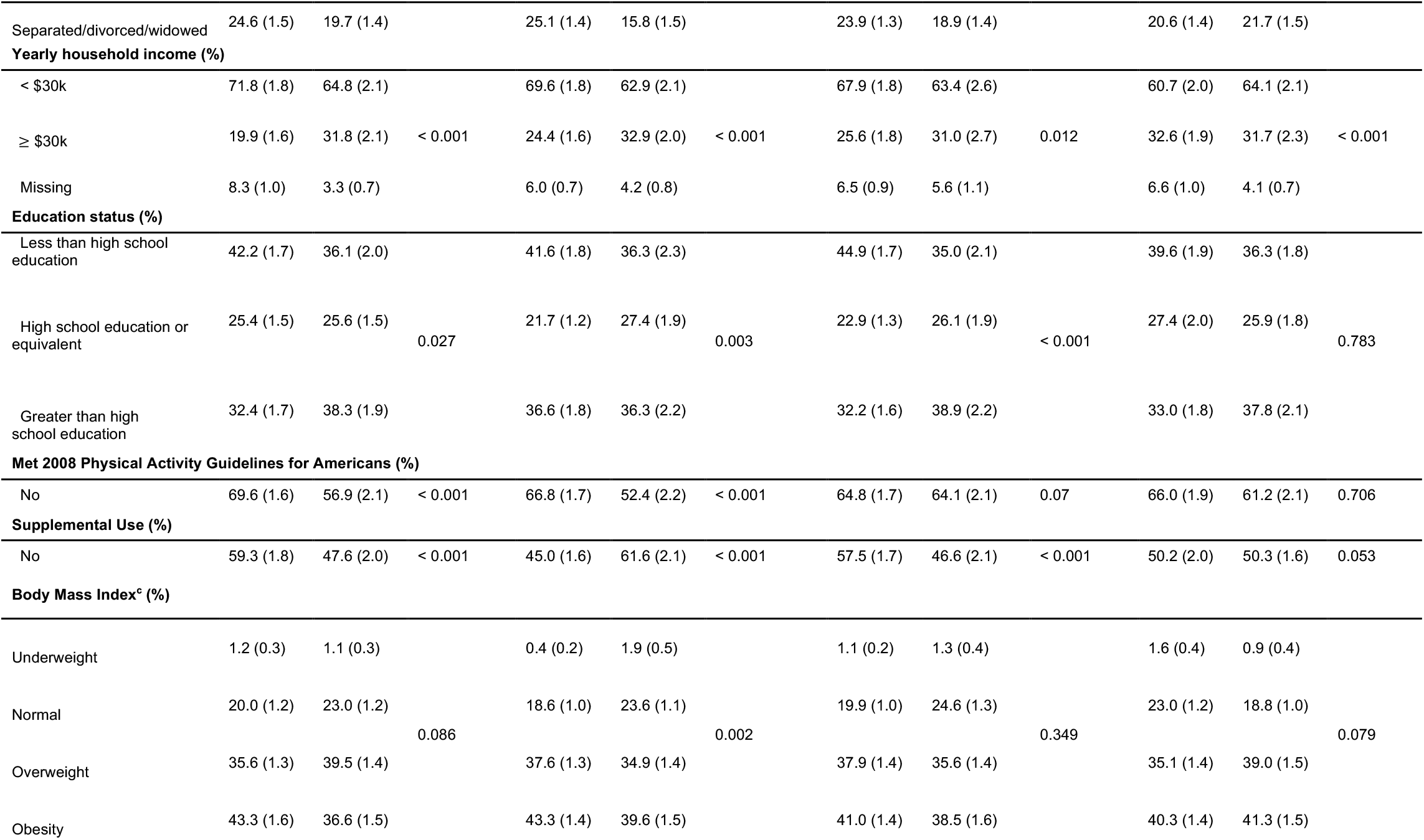
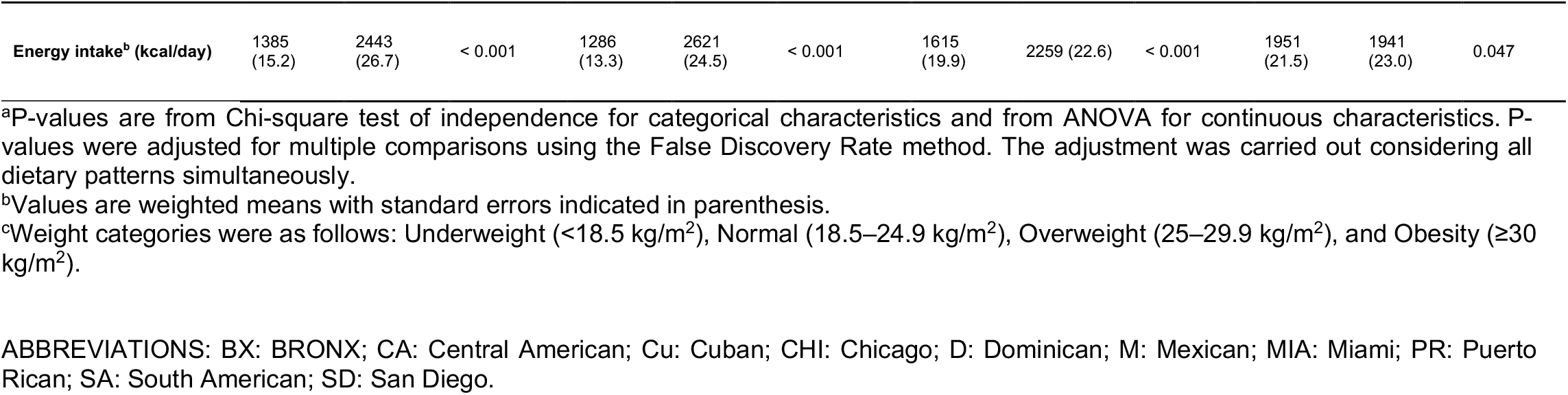
Socio-demographic and lifestyle characteristics (weighted percentages and standard error in parenthesis) for participants by shared quintile-based (Q1, Q5) categories. Hispanic Community Health Study/Study of Latinos, 2008-2011.

#### Ethnic background-site-specific (EBS) dietary patterns

The identified EBS-specific DPs were sparingly related to the selected socio-demographic and lifestyle factors. Age, sex, age of immigration, education, and use of supplements showed adjusted p-values<0.05 for a range of two (age of immigration) to seven (sex) DPs. The two DPs expressed by Chicago and San Diego individuals of Mexican background were significantly related to most (54% and 62%, respectively) of the selected variables in the same direction: top-consumers of these DPs were more likely to be younger, to be first generation immigrants, to have lived less than 10 years in the US, to be married/living with a partner, to report an income less than $30,000, and to have less than a high school education (**Supplementary Table 10**).

## Discussion

This paper explores the use of BMSFA to assess reproducibility of *a posteriori* nutrient-based DPs across the 12 EBS categories available in HCHS/SOL. Four DPs - *Plant-based foods, Processed foods, Dairy products*, and *Seafood* – were shared across all EBS categories and were therefore reproducible. One EBS-specific DP was additionally identified for each of the 12 categories. The 12 EBS-specific DPs represented variants of an animal profile associated with consumption of poultry, alcohol, and sometimes red meat. Most EBS-specific DPs showed additional similarities in profile and were further grouped into overarching DPs, i.e., an *<u>Animal vs. vegetable source</u>*, an *<u>Animal source only</u>*, and a *<u>Poultry vs. dairy products</u>* DPs. Shared DPs from BMSFA were similar to their counterparts identified in PCFA and FMSFA, but EBS-specific DPs from BMSFA showed more nuances than those from FMSFA.

The shared DPs capture traits common to all EBS-specific populations in HCHS/SOL, independently of background and site of recruitment, and are meant to reflect key aspects of the overall diet of Hispanic/Latino adults that reside in the US. These DPs target vegetable (*Plant-based* DP) and animal sources (*Dairy foods* and *Seafood* DPs) of foods and the fat component of foods, as derived from vegetable and animal sources (*Processed foods* DP). Despite the central role of cereals and meat in Hispanic/Latino diets (42-44), we have not identified separate shared DPs for grains (i.e., a DP loading high on starch and/or total sugars and/or fiber) or meat (i.e., a DP loading high on animal protein and/or cholesterol). Grains were rather eaten together with vegetables or meats; meat was also consumed in combination with vegetables or dairy products or was included in baked or fast-food products. However, except for *Processed foods* DP, no other combinations including grains or meat were prevalent enough to emerge as clear separate DPs common to all EBS-specific categories. The identified shared DPs and the absence of separate DPs for grains and meat likely reflect a combination of a general Hispanic/Latino culinary tradition (45) and acculturation to more US-American diets.

The 12 EBS-specific DPs summarize variants of that animal profile captured only in part by the shared DPs. Overarching DPs likely capture a combination of background-specific (e.g., culinary traditions, cultures, beliefs) (45) and site-specific (e.g., food access and environment) (46) factors. Curiously, every overarching DP presented with EBS-specific categories that differed in both background and site. On the other hand, at the lowest AHEI-2010 mean scores, individuals of Puerto Rican background from Bronx and Chicago showed strikingly different DPs; at the highest AHEI-2010 mean scores, similarities were found between DPs of Mexican background from San Diego and Chicago, but not with those from Bronx. This further suggests the need to conduct analyses at the cross-section of site and background levels and the importance of using *a posteriori* DPs, which revealed differences in background by site, unexpected from previous analyses based on the AHEI-2010 (47). A few previous studies identified *a posteriori* food-group-based DPs with factor or cluster analyses conducted on overall populations of Hispanics/Latinos living in the US or on multi-cultural populations from US, including Hispanics/Latinos and Non-Hispanic Whites and Blacks (23, 24, 48-51). Although we reinterpreted nutrient-based DPs with food groups, the comparison of our results with existing literature is fraught with differences in dietary collection tools, age and race/ethnicity of participants, sample size, type and preprocessing of input variables, and DP identification method. Earlier than HCHS/SOL, at least 4 studies identified *a posteriori* DPs of Hispanic/Latino adults residing in the US (48-51). We restricted our comparison to those deriving DPs on Hispanics/Latinos only (48, 50). Dietary patterns of Mexican American adults from the US National Health and Nutrition Examination Survey 2001/2002 were examined in one study (48), where 4 shared clusters, named *Poultry and alcohol, Milk and baked products, Traditional Mexican*, and *Meat*, were identified. The *Traditional Mexican* and the *Meat* DPs confirm that grains (tacos/tortillas in this case) are consumed in combination with legumes (*Traditional Mexican* DP) or (red) meat (*Meat* DP) and load high on two separate DPs. The *Poultry and alcohol* DP confirms that poultry is frequently consumed with alcohol, as in most of our EBS-specific DPs. Dairy products loaded high on a separate animal-source *Milk and baked products* DP; the high loadings on cakes, cookies, and pizza suggest similarities with our *Processed foods* DP. Another study (50) reported on DPs of 45-75 years old Puerto Rican adults from the Boston area and identified 3 FA-based DPs named *Meat, processed meat, and French fries, Rice, beans, and oils*, and *Sweets, sugary beverages, and dairy desserts*. The *Rice, beans, and oils* confirms the rice and legumes’ combination; it also reveals the major role of oils. The *Meat, processed meat, and French fries* and the *Sweets, sugary beverages, and dairy desserts* DPs share similarities with our *Processed foods* DP; however, while our DP likely reflects acculturation to the US-American diet, the previous DPs likely capture modern industrialized diets related to nutrition transition, as they were shown not to be related to acculturation (52).

Within HCHS/SOL, two previous papers derived *a posteriori* DPs using food items/groups at either ethnic background-specific (23) or EBS-specific (24) levels. Maldonado et al. (23) described one fully and 4 partially reproducible DPs derived using FA on 34 food groups from 24-hour recalls stratifying by 6 ethnic background-specific categories. Their fully reproducible *Burgers, Fries, & Soft drinks* overarching DP is fairly similar to our shared *Processed foods* DP. Consistent with our *Seafood* DP, Maldonado’s *Fish* DP (23) loaded high on fish and to less extent poultry, but it showed opposing loadings for poultry in Dominican background participants (23); we observed the same behavior of fish and poultry in the EBS-specific Dominican background – Bronx DP. Our *Dairy products* DP shares similarities with Maldonado’s *Egg & Cheese* DP (23); however, starchy vegetables and processed meats (in Dominican and Puerto Rican participants only) suggest partial overlapping with our *Processed foods* DP. Maldonado’s *White Rice, Beans, & Red Meats* partially overlaps with our *Plant-based foods* DP, which showed 7 additional vegetable and fruit groups other than beans and was more oriented towards consumption of whole grains compared to refined grains. Whereas Maldonado et al. derived DPs separately by ethnic background using standard FA (23), Stephenson et al. (24) used robust profile clustering to jointly classify individuals and 129 food propensity questionnaire items into global or local food patterns, based on 9 out of our 12 EBS categories. Stephenson’s global profiles favored a more frequent consumption of fruits, vegetables, poultry, and fish in *Global Profile g1*, and foods with oils, added sugars, and eggs in *Global Profile 2* (24). This indicates that *Global Profile 1* contains a mix of elements from our *Plant-based foods* and *Seafood* DPs, whereas *Global Profile 2* contains many elements of our *Processed foods* DP. Like our BMSFA results, chicken was commonly consumed across all EBS categories, but with different levels of consumption frequency.

The association between the *Processed foods* DP and younger age at immigration/more years spent in the US aligns with results suggesting dietary practice outcomes worsen with acculturation (e.g., (51)). Similarly, top-consumers of the *Plant-based foods* and *Seafood* DPs were less likely to have spent more than 10 years in the US. Currently, there is still little agreement on how acculturation should be measured (52-54) and regression models are needed to consider the more complex patterns including acculturation, socio-demographic factors, diet and/or health outcomes, likely within a mediation analysis approach (55).

The current analysis has strengths and limitations. The HCHS/SOL is the largest cohort of individuals of Hispanic and Latino background in the United States; the study design and probability sampling in urban areas with large ethnically diverse Hispanic and Latino populations provide adequate representation. The identification of shared and EBS-specific DPs via MSFA were carried out in a single step. Also, the introduction of prior distributions (which act like rotations) in BMSFA has provided the most straightforward interpretation of the EBS-specific DPs observed, compared to FMSFA. Improved interpretation of the EBS-specific DPs under the BMSFA is not at the expense of the shared DP interpretation, as there is equivalence with shared FMSFA-or PCFA-based DPs. Based on 24-hour recall data, our identified DPs may not represent the participant’s usual diet and may have failed to capture episodically consumed foods well. In general, self-reported dietary assessment tools are prone to measurement error and 24-hour recalls have been shown to underestimate total dietary intake (56). Systematic under-reporting of energy and protein intake was observed in a biomarker calibration study in HCHS/SOL that varied by ethnic background (57), which may explain some of the lower mean energy values in certain EBS categories.

In conclusion, this application of BMSFA within HCHS/SOL reveals shared DPs that clearly document dietary habits of Hispanics/Latinos who live in the US. Using single EBS-specific DPs grouped as overarching DPs showed additional similarities in animal food sources related to the cross-section between background and site of recruitment. Although our results on the association between shared DPs and acculturation suggest dietary practice outcomes worsen with acculturation, implications for potential preventive strategies require further specific efforts to assess the more complex patterns of acculturation, socio-demographic factors, diet and/or health outcomes.

## Supporting information

STROBE check-list

## Data Availability

Data sharing: Data used in this study are not publicly available, but can be obtained through an internal process of the study, based on a Data and Materials Distribution Agreement (DMDA). Supporting code to implement the methods performed in this study is publicly and freely available. In detail, we finalized an R package, named MSFA, to implement the methods presented in De Vito et al. [De Vito R, Bellio R, Trippa L, and Parmigiani G, 'Multi-study factor analysis' and 'Bayesian multi-study factor analysis for high-throughput biological data'] and their application in nutritional epidemiology. The package is available on GitHub: https://github.com/rdevito/MSFA

## Author contributions

RDV, BJS, and VE designed overall research plan; DS-A: assisted with the research plan and writing of the manuscript; AMS-R, JM, and MP provided advice on nutritional issues and a critical review of the manuscript; RDV performed most of the statistical analyses, with contributions from BS; RDV prepared all tables and figures except for Supplementary Figure 1; RDV assisted in writing the Methods section and provided critical review of the manuscript; BS assisted in writing the Results section and provided critical review of the manuscript; BP, GCC, SAB, DGF, MLD, LVH assisted with interpretation of results and provided critical review of the manuscript; VE wrote the paper; VE and RDV had primary responsibility for final content. AMS-R, MLD, and LVH were involved in obtaining the cohort funding; AMS-R, MLD, LVH, and DS-A were involved in conducting the study including dietary data collection. All authors reviewed and approved the final manuscript. The authors have declared no conflicts of interest.

## Data sharing

Data used in this study are not publicly available, but can be obtained through an internal process of the study, based on a Data and Materials Distribution Agreement (DMDA). Supporting code to implement the methods performed in this study is publicly and freely available. In detail, we finalized an R package, named *MSFA*, to implement the methods presented in De Vito et al. [De Vito R, Bellio R, Trippa L, and Parmigiani G, “Multi-study factor analysis” and “Bayesian multi-study factor analysis for high-throughput biological data”] and their application in nutritional epidemiology. The package is available on GitHub: https://github.com/rdevito/MSFA

## Acknowledgements

The authors thank the staff and participants of HCHS/SOL for their important contributions. Investigators website -http://www.cscc.unc.edu/hchs/.

