## Supplementary Material - text, table, figure for "Shared and ethnic background site-specific dietary patterns in the Hispanic Community Health Study/Study of Latinos (HCHS/SOL)"

De Vito R., Stephenson B., *et al.*

### Materials and methods

#### *Specification of variables and data preprocessing*

We carried out a sensitivity analysis to assess if: 1. discarding nutrient information based on one (reliable) recall only; 2. combining information from one and two recalls using the mean of the two available reliable recalls. In detail, we calculated the summary statistics for each nutrient in the one-recall and two-recalls scenarios, considering the single ethnic background and site (EBS)-specific combinations and the merged data (generated by combining data from all the EBS-specific combinations); we then considered percentage differences in median intakes between the two scenarios, overall and by EBS-specific combination. As only a few nutrients from the list [i.e., eicosapentaenoic acid (EPA), docosapentaenoic acid (DPA), vitamin C, alpha and beta carotene, beta-cryptoxanthin, and lycopene] differed substantially (30% or more) in median under the two scenarios on the merged dataset, we decided to use all the available information provided from one and two recalls.

Kernel density estimation plots were created to inspect ethnic background and site (EBS)-specific nutrient intake distributions and allowed to detect a bimodality behavior for some nutrients (i.e., sodium, potassium, phosphorus, manganese) in some EBS-specific combinations. Nutrient intakes were all log-transformed (base e) to improve adherence to the assumption of normality of the shared and EBS-specific factors, as well as of the EBS-specific errors, as required by MSFA.

***Statistical analysis***

Bayesian multi-study factor analysis (BMSFA) (1) was performed to describe the variance-covariance structure among 42 nutrients calculated using information from 24-hour recalls collected at baseline (2008-2011) from the Hispanic Community Health Study/Study of Latinos.

In the current analysis, the study level is represented by the cross-section of ethnic background and study site. We considered S = 12 EBS categories, each sharing the same P=42 nutrients. Study s has $n_s$ subjects contributing to the P-dimensional log-transformed and standardized vector, $x_{is}$, with i=1,…, $n_s$, s=1,…, S. The BMSFA estimates K shared factors common to all the studies and $J_s$ study-specific factors associated with each study. We can then assume that each variable in each study $x_{is}$ is decomposed as:

$$x_{is} = \Phi f_{is} + \Lambda_s l_{is} + e_{is} \quad i=1,\dots, n_s \qquad s=1,\dots, S,$$

where $f_{is}$ are the shared latent factors and Φ is its corresponding shared factor-loading matrix; $l_{is}$ are the study-specific latent factors, and $\Lambda_s$ are the corresponding study-specific factor-loading matrices, and, finally, the error term $e_{is}$ has a diagonal covariance matrix $\Psi_s$ = diag($\psi_{s1}$,…, $\psi_{sp}$). As a consequence, the covariance matrix in each study can be obtained in the following way:

$$\Sigma_s = \Phi\Phi^T + \Lambda_s\Lambda_s^T + \Psi_s = \Sigma_\Phi + \Sigma_{\Lambda_s} + \Psi_s$$

The BMSFA imposes a level of sparsity with an infinite Bayesian factor matrix estimation. The parameter estimation is carried out by a fast and efficient Gibbs Sampler (1), where two identifiability issues are solved: 1. label switching within each factor-loading matrix (switching a column with another within each factor-loading matrix); 2. invariance with respect to orthogonal transformation (factor loadings matrices are only identifiable up to orthogonal rotation). These two identifiability issues are solved in a one-step procedure using eigenvalue decomposition. Specifically, after running 20,000 iterations of Gibbs Sampler, we averaged $\Sigma_{\Phi}$ and consequently $\Sigma_{\Lambda_s}$. To derive our shared $\Phi$ and study-specific $\Lambda_s$ factor-loading matrices, we proceeded via the eigenvalue decomposition of $\Sigma_{\Phi}$ and $\Sigma_{\Lambda_s}$, as described in (1).

We chose the number of shared and study-specific factors by setting a threshold (5%) on the percentage of total variance explained by each factor, obtained by dividing each eigenvalue by the sum of eigenvalues.

Compared with the frequentist multi-study factor analysis (FMSFA) (2), the BMSFA offers two advantages. First, it provides a better-defined factor-loading structure via the sparsity obtained by the prior distribution. In particular, the prior adopted in the BMSFA shrinks small loadings to zero, as the action of the prior is similar to a factor rotation, like the varimax one (1). Second, it chooses the dimension of the shared and study-specific factors through a one-step procedure, whereas the FMSFA adopts a two-step approach (i.e., standard factor analysis techniques to identify the total number of factors plus the Akaike Information Criterion (AIC) and the Bayesian Information Criterion (BIC) to identify the number of shared factors).

### Results

#### *Factorability of the correlation matrices*

**Supplemental Table 1** provides results from checks of factorability for each EBS category and the overall combined population. The correlation matrix of the merged database was amenable to factor analysis: p-value for the Bartlett's test of sphericity (rejecting the null hypothesis that the correlation matrix is the identity matrix) $< 0.001$, overall measure of sampling adequacy (Kaiser-Meyer-Olkin measure) = 0.89, and individual measures of sampling adequacy ranging from 0.48 (manganese) to 0.97 (sodium), with only two nutrients with measures smaller than 0.60 (manganese and vitamin B12). The same message came from results at each EBS-specific category. Specifically, all p-values from Bartlett's test of sphericity were < 0.001, and the overall Kaiser-Meyer-Olkin statistic ranged from 0.83 (Mexican background – Bronx and South American background – Chicago) to 0.91 (Puerto Rican background – Bronx) across the 12 categories. This indicates that we had an adequate sample size relative to the number of nutrients within each EBS category. When looking at the individual measures of sampling adequacy, about 10-20 of the 42 selected nutrients were poorly captured in two EBS categories (12 for Cuban background – Miami, n=2241, and 17 for Mexican background – Bronx, n=205). Most of these nutrients also showed poor measures across other EBS categories (from 1 to 5 categories in total), suggesting that the following nutrients: medium-chain saturated fatty acid, arachidonic acid, EPA, DPA, manganese, vitamin B6, vitamin B12, retinol, and lycopene were more likely to be not well measured by the 24-hour recalls. However, this could happen differently across the EBS categories; for example, manganese showed values as low as 0.38 and 0.45 for participants of South American background from Chicago and Miami and values higher than 0.95 in most other EBS categories, likely suggesting a different dietary behavior across combinations.

#### *Identification of nutrient-based dietary patterns*

**Supplemental Tables 2** and **3** present single factor loadings and percentages of variance explained by each factor for the shared and EBS-specific DPs, respectively. The BMSFA estimated 4 shared dietary patterns (DPs), common to all EBS categories (explaining 62.5% of the total variance) and one EBS-specific DP for each of the 12 categories (explaining a variance ranging from 10.7% for Central American background – Miami to 14.4% for Puerto Rican background – Chicago).

#### *Internal reproducibility and consistency of the identified patterns*

**Supplemental Table 4** provides nutrient communalities for the DPs shared across all EBS categories (first column on the left) and for the EBS-specific DPs (remaining columns on the right). Communalities represent the proportion of each nutrient's variance that can be explained by the retained factors. They range between 0 and 1, with values ≥ 0.50 indicating that the factor analysis model is able to capture nutrient variability. Nutrient communalities were satisfactory, with most nutrient variances contributed by the retained factors at the shared and EBS-specific levels. Nutrients of note with low communalities included total sugars, vitamin C, beta carotene equivalent, lutein and zeaxanthin, and lycopene (values less than 0.34); these nutrients were not substantially captured by the factor analysis model at either the shared or the EBS-specific level. Furthermore, communalities for cholesterol and arachidonic acids improved from the overall analysis to the EBS-specific ones, thus suggesting that these nutrients were captured by the factor model at the EBS-specific level.

Standardized Cronbach's alphas for the shared factors were equal to 0.95, 0.94, 0.90, and 0.85, for the *Plant-based*, *Processed foods, Dairy products*, and *Seafood* DPs, respectively. Standardized Cronbach's alphas ranged from 0.80 (Mexican background – Bronx) to 0.91 (Mexican background – Chicago) for the EBS-specific DPs. Four EBS categories had standardized alphas reaching 0.90 (Chicago and San Diego participants of Mexican background, and Bronx and Chicago participants of Puerto Rican background). At the shared and EBS-specific levels, most of the standardized Cronbach's *alphas-when-item-deleted* were slightly lower than the corresponding alphas for the respective reduced factor, thus suggesting that deleting the nutrient under consideration did not substantially change the DP structure. The greater changes were observed for the *Seafood* DPs (standardized Cronbach's alpha=0.85), where deleting DPA and docosahexaenoic acid (DHA) made the Cronbach's *alphas-when-item-deleted* reach 0.78 and 0.79, respectively.

**Supplemental Tables 5**, **6**, **7**, and **8,** as well as **Figure 2,** present the comparison of DPs from BMSFA with those from FMSFA and a standard principal component factor analysis (PCFA). Following standard model selection procedures, the number of factors to retain was equal to 4 for both PCFA (i.e., the test of the hypothesis that 4 components are sufficient gave a root mean square of the residuals of 0.06 with the p-value of chi square < 0.001) and the FMSFA (i.e., AIC and BIC criteria). Moreover, the FMSFA confirmed the presence of one additional DP for each EBS category (i.e., each EBS-specific DP explained more than 5% of the total variance) as in BMSFA. Total variance explained by the 4 shared factors was similar to BMSFA (61.4% in the FMSFA and 58.5% in the PCFA). The order of the selected factors changed across the 3 solutions: the *Processed foods* DP ranked first in the PCFA, with the rest of the DPs being in the same order of BMSFA (**Supplemental Table 5**). The BMSFA-based shared DPs were equivalent to their counterparts from FMSFA (all congruence coefficients ≥ 0.97) and PCFA (all congruence coefficients ≥ 0.95) (**Figure 2**). Visual inspection of the factor-loading matrix of the shared DPs (**Supplemental Table 5**) suggested that BMSFA was more effective in: 1. shrinking moderately low (e.g., ~0.20) or high (e.g., 0.65) loadings (in absolute value) towards 0 or 1, respectively; 2. forcing nutrients close to being dominant on two DPs to become dominant on one DP only. These behaviors were more evident for nutrients in the *Plant-based* and *Dairy products* DPs. In detail, in the comparison with FMSFA, dominant and non-ubiquitous nutrients like vegetable protein, natural folate, soluble and insoluble fiber, as well as short- and medium-chain saturated fatty acids, calcium, and retinol received higher loadings from BMSFA, whereas vitamin B12 and vitamin D - which showed loadings smaller than 0.60 in FMSFA - well characterized the *Dairy products* DP in BMSFA. Similarly, in the comparison with PCFA, dominant nutrients like calcium and retinol showed even higher loadings, whereas small- and medium-chain saturated fatty acids became dominant nutrients in the *Dairy products* DP from BMSFA. In summary, besides calcium and retinol, which were present in all the three versions of the *Dairy products* DP, BMSFA identified 5 additional dominant nutrients, all with loadings ≥ 0.40 in the other solutions; two of them (i.e., vitamin B12 and vitamin D) were not dominant on that DP in any of the two additional solutions, two of them (i.e., small- and medium-chain saturated fatty acids) were dominant in the FMSFA solution but not in the PCFA, and riboflavin was dominant in the PCFA but loaded 0.60 on another DP (i.e., *Plant-based* DP) and 0.53 in the *Dairy products* DP in the FMSFA.

The FMSFA-specific DPs showed percentages of explained variances comparable to their BMSFA counterparts, ranging from 8.4% (Mexican background – Chicago) to 16.7% (Central American background – Bronx). However, they were all characterized by opposite signs targeting animal versus vegetable sources of foods. In 8 or more of these EBS-specific DPs, animal protein/cholesterol/arachidonic acid were opposed to vegetable protein/folate/soluble and insoluble fiber (**Supplementary Table 6**). Consistently with this interpretation, among the 66 (unique) CCs between pairs of FMSFA-based EBS-specific DPs, 9 (13.64%) suggested equivalence between DPs and 52 (78.79%) pointed to fair similarity, vs. one (equivalence) and 23 (fair similarity) for BMSFA-based EBS-specific DPs, respectively (**Supplementary Table 7**). This result confirms that FMSFA-based EBS-specific DPs showed fewer nuances than the corresponding BMSFA-based ones (fair similarity and equivalence: FMSFA 61 coefficients, 92.43%; BMSFA 24, 36.36%). The one-to-one comparison between FMSFA-based and BMSFA-based versions of the same EBS-specific DP supported this interpretation. Among the 12 EBS-specific DPs, only 4 (Bronx participants of Dominican and Central American background, Chicago and San Diego participants of Mexican background) showed fairly similar under the two approaches (**Supplementary Table 8**); those DPs opposed animal and vegetable sources in their BMSFA version too.

***Top consumers of shared dietary patterns by ethnic background-site category***

**Supplemental Table 9** provides weighted percentages (i.e., prevalence) of EBS-specific combinations in the top quintile category of each shared DP. Sixty-five percent of the EBS combinations showed percentages around the expected 20% (i.e., 18-22%), thus being well represented in the top quintile category of each shared DPs. Major deviations from the 18-22% interval were observed for the *Seafood* DP, with percentages as low as 13.1% (Mexican background – Bronx) and as high as 24.3% (South American background – Chicago). Notably, both EBS-specific categories compensated lower- or higher-than-expected percentages with corresponding percentages on the *Plant-based foods* DP (Mexican background – Bronx: 24.8%; South American background – Chicago: 17.4%), whereas the 16.8% of Mexican background – Chicago category on the *Seafood* DP was compensated by the 23.2% on the *Processed foods* DP. Compared to Mexican background participants from Chicago those from San Diego reached 23.7% on the Seafood DP. Participants of Puerto Rican background from Chicago showed a 16.5% prevalence on the Seafood DP.

***Socio-demographic and lifestyle factors associated with the Identified patterns***

*Ethnic background-site-specific (EBS) dietary patterns*

**Supplemental Table 10** shows socio-demographic and lifestyle characteristics of individuals by ethnic-background-site-specific tertile-based (Q1, Q3) categories. The identified EBS-specific DPs were generally unrelated to the selected socio-demographic and lifestyle factors. The two DPs expressed by Chicago and San Diego individuals of Mexican background were significantly related to most (54% and 62%, respectively) of the selected variables in the same direction: top-consumers of these DPs were more likely to be younger, immigrants of first generation, with less than 10 years of life in the US, to be married/living with a partner, to report an income less than $30,000, and to have less than a high school education level. These and other DPs were also sparingly related to other variables, including age, sex, age of immigration, education, and use of supplements, with adjusted p-values<0.05 for two (age of immigration) to seven (sex) DPs.

### Supplemental Tables

**Supplemental Table 1. Overall and individual measures of sampling adequacy[a], overall and by ethnic-background-site category. Hispanic Community Health Study/Study of Latinos, 2008-2011.**

| | Overall | BX D | BX CA | CHI CA | MIA CA | MIA Cu | BX M | CHI M | SD M | BX PR | CHI PR | CHI SA | MIA SA |
|---|---|---|---|---|---|---|---|---|---|---|---|---|---|
| **Overall measure** | 0.89 | 0.90 | 0.87 | 0.86 | 0.86 | 0.87 | 0.83 | 0.89 | 0.90 | 0.91 | 0.90 | 0.83 | 0.84 |
| **Individual measures** | | | | | | | | | | | | | |
| Animal Protein[b] (g) | 0.84 | 0.86 | 0.82 | 0.80 | 0.80 | 0.83 | 0.81 | 0.83 | 0.86 | 0.87 | 0.86 | 0.81 | 0.78 |
| Vegetable Protein (g) | 0.91 | 0.90 | 0.91 | 0.88 | 0.89 | 0.90 | 0.88 | 0.88 | 0.92 | 0.92 | 0.92 | 0.86 | 0.85 |
| Cholesterol (mg) | 0.90 | 0.89 | 0.85 | 0.80 | 0.76 | 0.80 | 0.76 | 0.88 | 0.90 | 0.89 | 0.87 | 0.80 | 0.70 |
| Short-chain saturated fatty acids (SCSFA) (g) | 0.79 | 0.78 | 0.77 | 0.74 | 0.63 | 0.74 | 0.76 | 0.81 | 0.81 | 0.85 | 0.82 | 0.62 | 0.76 |
| Medium-chain saturated fatty acids (MCSFA) (g) | 0.85 | 0.74 | 0.75 | 0.85 | 0.54 | 0.86 | 0.83 | 0.87 | 0.83 | 0.90 | 0.78 | 0.70 | 0.89 |
| Long-chain saturated fatty acids (LCSFA) (g) | 0.85 | 0.84 | 0.83 | 0.81 | 0.81 | 0.85 | 0.86 | 0.84 | 0.84 | 0.87 | 0.88 | 0.78 | 0.82 |
| Long-chain monounsaturated fatty acids (LCMFA) (g) | 0.86 | 0.86 | 0.84 | 0.82 | 0.83 | 0.87 | 0.83 | 0.84 | 0.84 | 0.87 | 0.85 | 0.81 | 0.82 |
| Linoleic Acid (g) | 0.90 | 0.90 | 0.84 | 0.89 | 0.89 | 0.93 | 0.84 | 0.89 | 0.86 | 0.91 | 0.86 | 0.89 | 0.85 |
| Linolenic Acid (g) | 0.95 | 0.96 | 0.88 | 0.91 | 0.94 | 0.93 | 0.90 | 0.96 | 0.94 | 0.94 | 0.97 | 0.91 | 0.90 |
| Arachidonic Acid (g) | 0.81 | 0.87 | 0.77 | 0.76 | 0.64 | 0.71 | 0.59 | 0.84 | 0.84 | 0.79 | 0.86 | 0.76 | 0.57 |
| Eicosapentaenoic Acid (EPA) (g) | 0.74 | 0.74 | 0.76 | 0.64 | 0.46 | 0.56 | 0.44 | 0.68 | 0.78 | 0.75 | 0.84 | 0.71 | 0.46 |
| Docosapentaenoic acid (DPA) (g) | 0.71 | 0.74 | 8.00 | 0.65 | 0.48 | 0.57 | 0.45 | 0.69 | 0.76 | 0.71 | 0.73 | 0.56 | 0.51 |
| Docosahexaenoic Acid (DHA) (g) | 0.79 | 0.71 | 0.72 | 0.80 | 0.88 | 0.82 | 0.84 | 0.75 | 0.77 | 0.69 | 0.74 | 0.71 | 0.82 |
| Total Trans Fatty Acids (g) | 0.93 | 0.95 | 0.89 | 0.85 | 0.83 | 0.89 | 0.75 | 0.90 | 0.92 | 0.94 | 0.90 | 0.88 | 0.85 |
| Total Sugars (g) | 0.96 | 0.95 | 0.86 | 0.94 | 0.93 | 0.92 | 0.93 | 0.96 | 0.95 | 0.96 | 0.95 | 0.88 | 0.94 |
| Starch (g) | 0.93 | 0.91 | 0.84 | 0.91 | 0.93 | 0.92 | 0.90 | 0.91 | 0.92 | 0.94 | 0.93 | 0.90 | 0.88 |
| Calcium (mg) | 0.93 | 0.92 | 0.91 | 0.89 | 0.89 | 0.90 | 0.89 | 0.92 | 0.94 | 0.92 | 0.93 | 0.86 | 0.89 |
| Phosphorus (mg) | 0.92 | 0.93 | 0.92 | 0.90 | 0.93 | 0.92 | 0.90 | 0.90 | 0.92 | 0.95 | 0.95 | 0.90 | 0.90 |
| Magnesium (mg) | 0.93 | 0.93 | 0.90 | 0.92 | 0.92 | 0.86 | 0.91 | 0.93 | 0.93 | 0.94 | 0.95 | 0.89 | 0.90 |

| | | | | | | | | | | | | | |
|---|---|---|---|---|---|---|---|---|---|---|---|---|---|
| Iron (mg) | 0.93 | 0.94 | 0.92 | 0.92 | 0.92 | 0.93 | 0.91 | 0.93 | 0.93 | 0.95 | 0.94 | 0.92 | 0.92 |
| Zinc (mg) | 0.93 | 0.92 | 0.90 | 0.89 | 0.91 | 0.91 | 0.90 | 0.93 | 0.92 | 0.95 | 0.88 | 0.85 | 0.88 |
| Copper (mg) | 0.77 | 0.84 | 0.80 | 0.78 | 0.72 | 0.78 | 0.73 | 0.81 | 0.70 | 0.89 | 0.88 | 0.67 | 0.82 |
| Selenium (mcg) | 0.93 | 0.96 | 0.92 | 0.91 | 0.90 | 0.93 | 0.79 | 0.91 | 0.93 | 0.94 | 0.92 | 0.91 | 0.88 |
| Sodium (mg) | 0.97 | 0.98 | 0.95 | 0.95 | 0.97 | 0.98 | 0.94 | 0.97 | 0.97 | 0.98 | 0.97 | 0.93 | 0.96 |
| Potassium (mg) | 0.91 | 0.89 | 0.87 | 0.90 | 0.92 | 0.88 | 0.91 | 0.93 | 0.93 | 0.92 | 0.92 | 0.91 | 0.87 |
| Manganese (mg) | 0.48 | 0.96 | 0.95 | 0.96 | 0.96 | 0.97 | 0.95 | 0.97 | 0.85 | 0.96 | 0.96 | 0.38 | 0.45 |
| Thiamin (vitamin B1) (mg) | 0.95 | 0.94 | 0.88 | 0.90 | 0.94 | 0.94 | 0.93 | 0.92 | 0.95 | 0.95 | 0.93 | 0.89 | 0.91 |
| Riboflavin (vitamin B2) (mg) | 0.92 | 0.91 | 0.88 | 0.91 | 0.91 | 0.89 | 0.87 | 0.93 | 0.94 | 0.92 | 0.90 | 0.88 | 0.88 |
| Niacin (vitamin B3) (mg) | 0.88 | 0.89 | 0.88 | 0.83 | 0.87 | 0.83 | 0.80 | 0.88 | 0.91 | 0.89 | 0.91 | 0.82 | 0.86 |
| Pantothenic Acid (vitamin B5) (mg) | 0.94 | 0.95 | 0.93 | 0.89 | 0.94 | 0.93 | 0.91 | 0.94 | 0.91 | 0.96 | 0.96 | 0.92 | 0.92 |
| Vitamin B6 (mg) | 0.70 | 0.93 | 0.90 | 0.59 | 0.91 | 0.50 | 0.54 | 0.92 | 0.93 | 0.91 | 0.89 | 0.90 | 0.91 |
| Vitamin B12 (mcg) | 0.50 | 0.76 | 0.69 | 0.36 | 0.91 | 0.40 | 0.51 | 0.91 | 0.90 | 0.83 | 0.82 | 0.76 | 0.83 |
| Natural Folate (mcg) | 0.91 | 0.90 | 0.86 | 0.90 | 0.90 | 0.91 | 0.86 | 0.88 | 0.90 | 0.92 | 0.92 | 0.83 | 0.84 |
| Vitamin C (mg) | 0.88 | 0.82 | 0.71 | 0.81 | 0.81 | 0.83 | 0.60 | 0.86 | 0.87 | 0.80 | 0.84 | 0.72 | 0.78 |
| Retinol (mcg) | 0.62 | 0.80 | 0.79 | 0.58 | 0.63 | 0.54 | 0.64 | 0.63 | 0.61 | 0.82 | 0.76 | 0.57 | 0.73 |
| Beta Carotene Equivalents (mcg) | 0.78 | 0.79 | 0.75 | 0.81 | 0.73 | 0.75 | 0.63 | 0.77 | 0.76 | 0.79 | 0.75 | 0.66 | 0.71 |
| Lutein + Zeaxanthin (mcg) | 0.81 | 0.85 | 0.68 | 0.62 | 0.76 | 0.71 | 0.66 | 0.78 | 0.71 | 0.87 | 0.74 | 0.65 | 0.73 |
| Lycopene (mcg) | 0.84 | 0.89 | 0.76 | 0.70 | 0.81 | 0.76 | 0.45 | 0.77 | 0.85 | 0.82 | 0.90 | 0.84 | 0.81 |
| Vitamin D (mcg) | 0.89 | 0.91 | 0.86 | 0.79 | 0.87 | 0.86 | 0.72 | 0.87 | 0.89 | 0.95 | 0.90 | 0.81 | 0.85 |
| Natural Alpha-Tocopherol (mg) | 0.90 | 0.92 | 0.91 | 0.89 | 0.89 | 0.89 | 0.88 | 0.90 | 0.93 | 0.92 | 0.89 | 0.73 | 0.81 |
| Soluble Dietary Fiber (g) | 0.93 | 0.95 | 0.93 | 0.91 | 0.90 | 0.90 | 0.87 | 0.93 | 0.92 | 0.96 | 0.92 | 0.88 | 0.93 |
| Insoluble Dietary Fiber (g) | 0.95 | 0.95 | 0.93 | 0.93 | 0.91 | 0.94 | 0.87 | 0.94 | 0.94 | 0.95 | 0.95 | 0.88 | 0.93 |

[a] Overall and individual measures of sampling adequacy range between 0 and 1, with values > 0.60 indicating a satisfactory size. [b] Units of the nutrients indicated their original scale.
ABBREVIATIONS: BX: BRONX; CA: Central American; Cu: Cuban; CHI: Chicago; D: Dominican; M: Mexican; MIA: Miami; PR: Puerto Rican; SA: South American; SD: San Diego.

**Supplemental Table 2. Factor-loading matrix and explained variances[a] for the 4 shared dietary patterns identified by the Bayesian multi-study factor analysis. Hispanic Community Health Study/Study of Latinos, 2008-2011.**

| Nutrient | *Plant-based foods* | *Processed foods* | *Dairy products* | *Seafood* |
|---|---|---|---|---|
| Animal Protein[b,c] (g) | 0.27 | 0.43 | 0.48 | 0.27 |
| Vegetable Protein (g) | **0.80** | 0.34 | 0.09 | -0.02 |
| Cholesterol (mg) | 0.12 | 0.50 | 0.42 | 0.25 |
| Short-chain saturated fatty acids (SCSFA) (g) | -0.04 | 0.25 | **0.79** | -0.09 |
| Medium-chain saturated fatty acids (MCSFA) (g) | -0.02 | 0.27 | **0.71** | -0.08 |
| Long-chain saturated fatty acids (LCSFA) (g) | 0.15 | **0.78** | 0.53 | -0.03 |
| Long-chain monounsaturated fatty acids (LCMFA) (g) | 0.29 | **0.92** | 0.18 | 0.04 |
| Linoleic Acid (g) | 0.30 | **0.86** | 0.02 | 0.06 |
| Linolenic Acid (g) | 0.31 | **0.70** | 0.13 | 0.07 |
| Arachidonic Acid (g) | 0.18 | 0.39 | 0.14 | 0.43 |
| Eicosapentaenoic Acid (EPA) (g) | 0.06 | 0.00 | 0.05 | **0.88** |
| Docosapentaenoic acid (DPA) (g) | 0.12 | 0.06 | 0.01 | **0.84** |
| Docosahexaenoic Acid (DHA) (g) | 0.09 | 0.07 | 0.06 | **0.97** |
| Total Trans Fatty Acids (g) | 0.04 | **0.60** | 0.36 | -0.02 |
| Total Sugars (g) | 0.31 | 0.25 | 0.38 | -0.03 |
| Starch (g) | 0.57 | 0.50 | 0.14 | -0.04 |
| Calcium (mg) | 0.41 | 0.12 | **0.76** | 0.00 |
| Phosphorus (mg) | **0.63** | 0.37 | 0.57 | 0.18 |
| Magnesium (mg) | **0.85** | 0.23 | 0.28 | 0.10 |
| Iron (mg) | **0.71** | 0.32 | 0.29 | 0.02 |
| Zinc (mg) | **0.61** | 0.39 | 0.42 | 0.02 |
| Copper (mg) | **0.77** | 0.25 | 0.16 | 0.10 |

| | | | | |
|---|---|---|---|---|
| Selenium (mcg) | 0.48 | 0.54 | 0.33 | 0.30 |
| Sodium (mg) | 0.44 | 0.59 | 0.26 | 0.09 |
| Potassium (mg) | **0.78** | 0.23 | 0.33 | 0.12 |
| Manganese (mg) | **0.75** | 0.20 | 0.09 | 0.04 |
| Thiamin (vitamin B1) (mg) | **0.63** | 0.43 | 0.35 | -0.02 |
| Riboflavin (vitamin B2) (mg) | 0.50 | 0.24 | **0.69** | 0.04 |
| Niacin (vitamin B3) (mg) | **0.60** | 0.39 | 0.24 | 0.18 |
| Pantothenic Acid (vitamin B5) (mg) | **0.63** | 0.26 | 0.45 | 0.17 |
| Vitamin B6 (mg) | **0.73** | 0.22 | 0.28 | 0.13 |
| Vitamin B12 (mcg) | 0.34 | 0.12 | **0.60** | 0.23 |
| Natural Folate (mcg) | **0.74** | 0.19 | 0.11 | 0.06 |
| Vitamin C (mg) | 0.47 | 0.02 | 0.08 | 0.07 |
| Retinol (mcg) | 0.20 | 0.06 | **0.70** | 0.06 |
| Beta Carotene Equivalents (mcg) | 0.37 | -0.02 | 0.03 | 0.10 |
| Lutein + Zeaxanthin (mcg) | 0.39 | 0.08 | 0.09 | 0.13 |
| Lycopene (mcg) | 0.19 | 0.15 | 0.05 | -0.01 |
| Vitamin D (mcg) | 0.23 | 0.03 | **0.66** | 0.31 |
| Natural Alpha-Tocopherol (mg) | 0.54 | **0.60** | 0.06 | 0.16 |
| Soluble Dietary Fiber (g) | **0.68** | 0.15 | 0.12 | 0.00 |
| Insoluble Dietary Fiber (g) | **0.84** | 0.14 | 0.03 | 0.01 |
| Proportion of variance explained (%) | 25.1 | 15.3 | 14.3 | 7.7 |
| Cumulative variance explained (%) | 25.1 | 40.4 | 54.7 | 62.4 |

[a] Estimated from the Bayesian multi-study factor analysis carried out on 42 nutrients. The magnitude of each loading measures the importance of the corresponding nutrient to the factor. [b] Loadings ≥0.60 define the dominant nutrients for each factor and were shown in bold typeface. [c] Units of the nutrients indicated their original scale, but loadings were derived from log transformed and standardized nutrient intakes entered into the Bayesian multi-study factor analysis model.

**Supplemental Table 3. Factor-loading matrix and explained variances[a] for the 12 ethnic-background-site-specific dietary patterns identified by the Bayesian multi-study factor analysis. Hispanic Community Health Study/Study of Latinos, 2008-2011.**

| Nutrient | BX D | BX CA | CHI CA | MIA CA | MIA Cu | BX M | CHI M | SD M | BX PR | CHI PR | CHI SA | MIA SA |
|---|---|---|---|---|---|---|---|---|---|---|---|---|
| Animal Protein[b,c] (g) | **0.47** | **0.40** | **0.61** | **0.48** | **0.55** | **0.37** | **0.34** | 0.22 | **0.50** | 0.05 | **0.44** | **0.55** |
| Vegetable Protein (g) | -0.11 | 0.02 | -0.21 | -0.18 | -0.07 | -0.28 | -0.21 | -0.19 | -0.20 | -0.24 | -0.11 | -0.21 |
| Cholesterol (mg) | **0.33** | 0.23 | **0.34** | **0.30** | **0.40** | 0.22 | 0.17 | 0.06 | **0.36** | 0.13 | 0.29 | **0.46** |
| Short-chain saturated fatty acids (SCSFA) (g) | -0.22 | **-0.37** | -0.23 | **-0.36** | -0.28 | **-0.38** | -0.11 | -0.18 | -0.25 | 0.04 | -0.26 | -0.20 |
| Medium-chain saturated fatty acids (MCSFA) (g) | -0.22 | **-0.41** | -0.24 | **-0.35** | **-0.30** | **-0.36** | -0.11 | -0.20 | -0.28 | 0.00 | -0.23 | -0.23 |
| Long-chain saturated fatty acids (LCSFA) (g) | 0.00 | -0.16 | 0.05 | -0.07 | 0.06 | -0.10 | 0.03 | -0.09 | 0.00 | 0.01 | 0.09 | 0.09 |
| Long-chain monounsaturated fatty acids (LCMFA) (g) | -0.01 | -0.06 | 0.00 | 0.00 | 0.09 | -0.06 | 0.05 | -0.09 | 0.00 | 0.05 | 0.10 | 0.09 |
| Linoleic Acid (g) | 0.01 | -0.05 | -0.09 | 0.00 | 0.05 | -0.10 | 0.01 | -0.09 | -0.06 | 0.07 | 0.09 | 0.02 |
| Linolenic Acid (g) | -0.06 | -0.19 | -0.12 | -0.11 | -0.05 | -0.15 | -0.01 | -0.16 | -0.17 | 0.07 | -0.04 | -0.06 |
| Arachidonic Acid (g) | **0.44** | **0.43** | **0.44** | **0.41** | **0.49** | **0.40** | 0.24 | 0.12 | **0.51** | 0.02 | **0.34** | **0.57** |
| Eicosapentaenoic Acid (EPA) (g) | -0.16 | -0.05 | -0.11 | -0.19 | -0.16 | -0.09 | -0.09 | -0.09 | -0.16 | -0.10 | -0.11 | -0.22 |
| Docosapentaenoic acid (DPA) (g) | 0.13 | 0.23 | 0.16 | 0.07 | 0.10 | 0.09 | 0.04 | 0.00 | 0.12 | -0.09 | 0.00 | 0.13 |
| Docosahexaenoic Acid (DHA) (g) | -0.04 | 0.09 | -0.02 | -0.05 | -0.04 | 0.03 | -0.03 | -0.06 | -0.01 | -0.09 | -0.05 | -0.07 |
| Total Trans Fatty Acids (g) | 0.06 | -0.10 | 0.01 | -0.08 | -0.03 | -0.04 | 0.09 | 0.00 | -0.03 | -0.06 | 0.06 | 0.08 |
| Total Sugars (g) | -0.17 | -0.17 | -0.06 | -0.10 | -0.18 | 0.00 | 0.05 | -0.03 | -0.17 | 0.00 | -0.16 | -0.11 |
| Starch (g) | -0.05 | 0.02 | -0.13 | -0.02 | -0.01 | -0.23 | -0.10 | 0.01 | -0.19 | **-0.31** | 0.14 | -0.10 |
| Calcium (mg) | -0.14 | -0.14 | -0.16 | -0.16 | -0.11 | -0.14 | -0.09 | -0.10 | -0.22 | 0.01 | -0.16 | -0.24 |
| Phosphorus (mg) | 0.09 | 0.11 | 0.11 | 0.05 | 0.17 | -0.07 | -0.11 | -0.05 | 0.06 | -0.02 | 0.09 | 0.05 |
| Magnesium (mg) | -0.16 | -0.06 | 0.00 | -0.10 | 0.02 | -0.11 | -0.21 | -0.19 | -0.07 | 0.11 | -0.09 | -0.12 |
| Iron (mg) | 0.16 | 0.23 | -0.02 | 0.18 | 0.15 | 0.10 | 0.20 | **0.31** | 0.03 | **-0.36** | 0.22 | 0.02 |
| Zinc (mg) | **0.30** | 0.29 | 0.28 | **0.33** | **0.41** | 0.20 | 0.17 | 0.21 | 0.26 | -0.11 | **0.32** | 0.29 |
| Copper (mg) | -0.07 | 0.04 | -0.02 | 0.07 | 0.08 | 0.01 | -0.09 | -0.13 | -0.08 | -0.03 | -0.01 | -0.06 |

| | | | | | | | | | | | | |
|---|---|---|---|---|---|---|---|---|---|---|---|---|
| Selenium (mcg) | **0.31** | **0.30** | 0.27 | 0.29 | **0.35** | 0.23 | 0.21 | 0.11 | 0.23 | -0.18 | 0.30 | 0.28 |
| Sodium (mg) | 0.14 | 0.01 | 0.08 | 0.12 | 0.18 | -0.04 | 0.15 | 0.04 | 0.05 | -0.18 | 0.16 | 0.12 |
| Potassium (mg) | -0.21 | -0.13 | 0.03 | -0.09 | 0.07 | -0.02 | -0.11 | -0.20 | -0.03 | 0.23 | -0.11 | 0.02 |
| Manganese (mg) | -0.09 | -0.01 | -0.14 | -0.05 | -0.02 | -0.10 | -0.09 | -0.10 | -0.13 | -0.09 | -0.12 | -0.06 |
| Thiamin (vitamin B1) (mg) | 0.09 | 0.12 | -0.09 | 0.09 | 0.15 | 0.02 | 0.18 | 0.22 | -0.03 | **-0.36** | 0.12 | 0.02 |
| Riboflavin (vitamin B2) (mg) | 0.07 | 0.05 | -0.04 | 0.16 | 0.15 | 0.13 | 0.26 | 0.26 | 0.00 | -0.13 | 0.08 | 0.01 |
| Niacin (vitamin B3) (mg) | **0.38** | **0.39** | **0.34** | **0.41** | **0.37** | **0.37** | **0.48** | **0.43** | 0.29 | -0.23 | **0.45** | 0.27 |
| Pantothenic Acid (vitamin B5) (mg) | 0.15 | 0.23 | 0.11 | 0.24 | 0.25 | 0.21 | 0.24 | 0.11 | 0.11 | 0.06 | 0.18 | 0.19 |
| Vitamin B6 (mg) | 0.16 | 0.24 | **0.30** | 0.29 | **0.32** | **0.36** | **0.30** | **0.32** | 0.20 | -0.03 | 0.25 | 0.21 |
| Vitamin B12 (mcg) | 0.15 | 0.07 | 0.14 | **0.33** | 0.28 | **0.34** | **0.36** | **0.37** | 0.10 | -0.09 | 0.26 | 0.02 |
| Natural Folate (mcg) | -0.29 | -0.25 | -0.13 | -0.25 | -0.09 | -0.21 | -0.29 | **-0.36** | -0.16 | 0.21 | **-0.35** | -0.11 |
| Vitamin C (mg) | **-0.33** | -0.17 | 0.00 | -0.08 | -0.05 | 0.09 | 0.03 | -0.15 | -0.14 | 0.23 | -0.22 | -0.01 |
| Retinol (mcg) | 0.00 | -0.03 | -0.25 | 0.00 | -0.07 | 0.01 | 0.12 | 0.16 | -0.12 | -0.06 | 0.07 | -0.17 |
| Beta Carotene Equivalents (mcg) | **-0.32** | **-0.32** | 0.07 | -0.12 | -0.07 | 0.00 | 0.02 | -0.24 | -0.15 | **0.31** | -0.15 | 0.01 |
| Lutein + Zeaxanthin (mcg) | -0.25 | -0.22 | -0.01 | -0.06 | -0.02 | 0.03 | 0.04 | -0.21 | -0.13 | 0.27 | -0.23 | 0.01 |
| Lycopene (mcg) | -0.14 | -0.07 | 0.20 | 0.01 | -0.06 | 0.03 | 0.01 | -0.10 | -0.08 | 0.07 | -0.06 | -0.08 |
| Vitamin D (mcg) | -0.07 | -0.05 | -0.17 | -0.04 | -0.02 | 0.00 | 0.07 | 0.11 | -0.08 | 0.01 | -0.01 | -0.22 |
| Natural Alpha-Tocopherol (mg) | -0.22 | -0.20 | -0.06 | -0.12 | -0.02 | -0.10 | -0.07 | -0.25 | -0.16 | 0.13 | -0.18 | -0.03 |
| Soluble Dietary Fiber (g) | **-0.35** | **-0.30** | -0.19 | **-0.35** | -0.18 | -0.26 | **-0.39** | **-0.37** | -0.27 | 0.13 | **-0.34** | -0.14 |
| Insoluble Dietary Fiber (g) | **-0.37** | -0.29 | -0.14 | **-0.32** | -0.15 | -0.21 | **-0.39** | **-0.33** | -0.22 | 0.15 | **-0.37** | -0.24 |
| Proportion of variance explained (%) | 11.7 | 11.9 | 12.8 | 10.7 | 11.1 | 11.6 | 13.3 | 11.4 | 12.2 | 14.4 | 11.2 | 12.7 |
| Cumulative variance explained (%) | 74.2 | 74.4 | 75.3 | 73.2 | 73.6 | 74.1 | 75.8 | 73.9 | 74.7 | 76.9 | 73.7 | 75.2 |

[a]Estimated from a Bayesian multi-study factor analysis carried out on forty-two nutrients. The magnitude of each loading measures the importance of the corresponding nutrient to the factor. [b]Loadings ≥0.3 in absolute value define the dominant nutrients for each factor and were shown in bold typeface. [c]Units of the nutrients indicated their original scale, but loadings were derived from log-transformed and standardized nutrient intakes entered into the Bayesian multi-study factor analysis model.

ABBREVIATIONS: BX: BRONX; CA: Central American; Cu: Cuban; CHI: Chicago; D: Dominican; M: Mexican; MIA: Miami; PR: Puerto Rican; SA: South American; SD: San Diego.

**Supplemental Table 4. Nutrient communalities[a] for the shared and ethnic-background-site-specific dietary patterns identified with the Bayesian multi-study factor analysis. Hispanic Community Health Study/Study of Latinos, 2008-2011.**

| Nutrient | Overall | BX D | BX CA | CHI CA | MIA CA | MIA Cu | BX M | CHI M | SD M | BX PR | CHI PR | CHI SA | MIA SA |
|---|---|---|---|---|---|---|---|---|---|---|---|---|---|
| Animal Protein[b] (g) | 0.56 | 0.78 | 0.71 | 0.93 | 0.78 | 0.87 | 0.70 | 0.67 | 0.60 | 0.81 | 0.56 | 0.75 | 0.86 |
| Vegetable Protein (g) | 0.76 | 0.77 | 0.76 | 0.80 | 0.79 | 0.77 | 0.84 | 0.80 | 0.80 | 0.80 | 0.82 | 0.77 | 0.8 |
| Cholesterol (mg) | 0.50 | 0.62 | 0.56 | 0.62 | 0.59 | 0.66 | 0.55 | 0.53 | 0.51 | 0.63 | 0.52 | 0.59 | 0.71 |
| Short-chain saturated fatty acids (SCSFA) (g) | 0.70 | 0.75 | 0.83 | 0.75 | 0.83 | 0.78 | 0.84 | 0.71 | 0.73 | 0.76 | 0.70 | 0.77 | 0.74 |
| Medium-chain saturated fatty acids (MCSFA) (g) | 0.58 | 0.63 | 0.75 | 0.64 | 0.71 | 0.67 | 0.71 | 0.59 | 0.62 | 0.66 | 0.58 | 0.64 | 0.63 |
| Long-chain saturated fatty acids (LCSFA) (g) | 0.90 | 0.90 | 0.93 | 0.91 | 0.91 | 0.91 | 0.92 | 0.91 | 0.91 | 0.90 | 0.90 | 0.91 | 0.91 |
| Long-chain monounsaturated fatty acids (LCMFA) (g) | 0.96 | 0.96 | 0.96 | 0.96 | 0.96 | 0.97 | 0.96 | 0.96 | 0.97 | 0.96 | 0.96 | 0.97 | 0.97 |
| Linoleic Acid (g) | 0.83 | 0.83 | 0.83 | 0.84 | 0.83 | 0.83 | 0.84 | 0.83 | 0.84 | 0.83 | 0.83 | 0.84 | 0.83 |
| Linolenic Acid (g) | 0.61 | 0.62 | 0.65 | 0.62 | 0.62 | 0.61 | 0.63 | 0.61 | 0.64 | 0.64 | 0.62 | 0.61 | 0.62 |
| Arachidonic Acid (g) | 0.39 | 0.58 | 0.57 | 0.58 | 0.56 | 0.62 | 0.55 | 0.45 | 0.40 | 0.65 | 0.39 | 0.51 | 0.71 |
| Eicosapentaenoic Acid (EPA) (g) | 0.78 | 0.80 | 0.78 | 0.79 | 0.81 | 0.80 | 0.79 | 0.79 | 0.79 | 0.80 | 0.79 | 0.79 | 0.83 |
| Docosapentaenoic acid (DPA) (g) | 0.72 | 0.74 | 0.78 | 0.75 | 0.73 | 0.73 | 0.73 | 0.72 | 0.72 | 0.74 | 0.73 | 0.72 | 0.74 |
| Docosahexaenoic Acid (DHA) (g) | 0.96 | 0.97 | 0.97 | 0.96 | 0.97 | 0.97 | 0.97 | 0.96 | 0.97 | 0.96 | 0.97 | 0.97 | 0.97 |
| Total Trans Fatty Acids (g) | 0.49 | 0.49 | 0.50 | 0.49 | 0.49 | 0.49 | 0.49 | 0.50 | 0.49 | 0.49 | 0.49 | 0.49 | 0.49 |
| Total Sugars (g) | 0.30 | 0.33 | 0.33 | 0.31 | 0.31 | 0.34 | 0.30 | 0.30 | 0.30 | 0.33 | 0.30 | 0.33 | 0.31 |
| Starch (g) | 0.59 | 0.60 | 0.59 | 0.61 | 0.59 | 0.59 | 0.65 | 0.60 | 0.59 | 0.63 | 0.69 | 0.61 | 0.6 |
| Calcium (mg) | 0.76 | 0.78 | 0.78 | 0.79 | 0.79 | 0.77 | 0.78 | 0.77 | 0.77 | 0.81 | 0.76 | 0.79 | 0.82 |
| Phosphorus (mg) | 0.88 | 0.89 | 0.89 | 0.89 | 0.88 | 0.91 | 0.89 | 0.89 | 0.89 | 0.89 | 0.88 | 0.89 | 0.89 |
| Magnesium (mg) | 0.87 | 0.89 | 0.87 | 0.87 | 0.88 | 0.87 | 0.88 | 0.91 | 0.90 | 0.87 | 0.88 | 0.87 | 0.88 |
| Iron (mg) | 0.69 | 0.72 | 0.75 | 0.69 | 0.73 | 0.72 | 0.70 | 0.73 | 0.79 | 0.70 | 0.82 | 0.74 | 0.69 |
| Zinc (mg) | 0.70 | 0.79 | 0.79 | 0.78 | 0.81 | 0.87 | 0.75 | 0.73 | 0.75 | 0.77 | 0.72 | 0.81 | 0.79 |
| Copper (mg) | 0.69 | 0.69 | 0.69 | 0.69 | 0.69 | 0.69 | 0.69 | 0.69 | 0.70 | 0.69 | 0.69 | 0.69 | 0.69 |

| | | | | | | | | | | | | | |
|---|---|---|---|---|---|---|---|---|---|---|---|---|---|
| Selenium (mcg) | 0.72 | 0.82 | 0.81 | 0.80 | 0.81 | 0.85 | 0.78 | 0.77 | 0.74 | 0.78 | 0.76 | 0.81 | 0.8 |
| Sodium (mg) | 0.62 | 0.64 | 0.62 | 0.62 | 0.63 | 0.65 | 0.62 | 0.64 | 0.62 | 0.62 | 0.65 | 0.64 | 0.63 |
| Potassium (mg) | 0.79 | 0.83 | 0.81 | 0.79 | 0.80 | 0.79 | 0.79 | 0.80 | 0.83 | 0.79 | 0.84 | 0.8 | 0.79 |
| Manganese (mg) | 0.62 | 0.63 | 0.62 | 0.64 | 0.62 | 0.62 | 0.63 | 0.63 | 0.63 | 0.63 | 0.63 | 0.63 | 0.62 |
| Thiamin (vitamin B1) (mg) | 0.71 | 0.71 | 0.72 | 0.72 | 0.72 | 0.73 | 0.71 | 0.74 | 0.75 | 0.71 | 0.84 | 0.72 | 0.71 |
| Riboflavin (vitamin B2) (mg) | 0.78 | 0.79 | 0.78 | 0.78 | 0.81 | 0.80 | 0.80 | 0.85 | 0.85 | 0.78 | 0.80 | 0.79 | 0.78 |
| Niacin (vitamin B3) (mg) | 0.60 | 0.74 | 0.75 | 0.71 | 0.77 | 0.73 | 0.73 | 0.83 | 0.78 | 0.68 | 0.65 | 0.8 | 0.67 |
| Pantothenic Acid (vitamin B5) (mg) | 0.70 | 0.72 | 0.75 | 0.71 | 0.76 | 0.76 | 0.74 | 0.76 | 0.71 | 0.71 | 0.70 | 0.73 | 0.74 |
| Vitamin B6 (mg) | 0.68 | 0.70 | 0.73 | 0.77 | 0.76 | 0.78 | 0.81 | 0.77 | 0.78 | 0.72 | 0.68 | 0.74 | 0.72 |
| Vitamin B12 (mcg) | 0.54 | 0.57 | 0.55 | 0.56 | 0.65 | 0.62 | 0.66 | 0.67 | 0.68 | 0.56 | 0.55 | 0.61 | 0.55 |
| Natural Folate (mcg) | 0.60 | 0.68 | 0.66 | 0.61 | 0.66 | 0.60 | 0.64 | 0.68 | 0.73 | 0.62 | 0.64 | 0.72 | 0.61 |
| Vitamin C (mg) | 0.23 | 0.34 | 0.26 | 0.23 | 0.24 | 0.23 | 0.24 | 0.23 | 0.25 | 0.25 | 0.28 | 0.28 | 0.23 |
| Retinol (mcg) | 0.54 | 0.54 | 0.54 | 0.61 | 0.54 | 0.55 | 0.54 | 0.56 | 0.57 | 0.56 | 0.55 | 0.55 | 0.57 |
| Beta Carotene Equivalents (mcg) | 0.15 | 0.25 | 0.25 | 0.15 | 0.16 | 0.15 | 0.15 | 0.15 | 0.20 | 0.17 | 0.24 | 0.17 | 0.15 |
| Lutein + Zeaxanthin (mcg) | 0.18 | 0.24 | 0.23 | 0.18 | 0.18 | 0.18 | 0.18 | 0.18 | 0.23 | 0.20 | 0.25 | 0.24 | 0.18 |
| Lycopene (mcg) | 0.06 | 0.08 | 0.07 | 0.10 | 0.06 | 0.07 | 0.06 | 0.06 | 0.07 | 0.07 | 0.07 | 0.07 | 0.07 |
| Vitamin D (mcg) | 0.58 | 0.58 | 0.58 | 0.61 | 0.58 | 0.58 | 0.58 | 0.58 | 0.59 | 0.58 | 0.58 | 0.58 | 0.62 |
| Natural Alpha-Tocopherol (mg) | 0.69 | 0.74 | 0.73 | 0.69 | 0.70 | 0.69 | 0.70 | 0.69 | 0.75 | 0.71 | 0.70 | 0.72 | 0.69 |
| Soluble Dietary Fiber (g) | 0.50 | 0.62 | 0.59 | 0.53 | 0.62 | 0.53 | 0.57 | 0.65 | 0.63 | 0.57 | 0.51 | 0.62 | 0.52 |
| Insoluble Dietary Fiber (g) | 0.72 | 0.86 | 0.81 | 0.74 | 0.83 | 0.75 | 0.77 | 0.87 | 0.83 | 0.77 | 0.75 | 0.86 | 0.78 |

[a]Nutrient communalities range between 0 and 1 with values ≥ 0.50 indicating that the factor analysis model (at the overall or at the ethnic-background-site-specific levels) is able to capture nutrient variability. The higher the communality the more the factor analysis model is able to capture nutrient variability. [b] Units of the nutrients indicated their original scale.

ABBREVIATIONS: BX: BRONX; CA: Central American; Cu: Cuban; CHI: Chicago; D: Dominican; M: Mexican; MIA: Miami; PR: Puerto Rican; SA: South American; SD: San Diego.

**Supplemental Table 5. Factor-loading matrix and explained variances[a] for the 4 dietary patterns identified with the frequentist multi-study factor analysis (shared patterns only) and with the principal component factor analysis. Hispanic Community Health Study/Study of Latinos, 2008-2011.**

| | **Frequentist multi-study factor analysis (MSFA)** | | | | **Principal component factor analysis (PCFA)** | | | |
|---|---|---|---|---|---|---|---|---|
| **Nutrient** | ***Plant-based foods*** | ***Processed foods*** | ***Dairy products*** | ***Seafood*** | ***Processed foods*** | ***Plant-based foods*** | ***Dairy products*** | ***Seafood*** |
| Animal Protein[b,c] (g) | 0.39 | 0.53 | 0.23 | 0.23 | **0.70** | 0.08 | 0.32 | 0.35 |
| Vegetable Protein (g) | **0.73** | 0.36 | 0.10 | -0.01 | 0.43 | **0.77** | 0.11 | -0.06 |
| Cholesterol (mg) | 0.20 | 0.57 | 0.25 | 0.22 | **0.62** | -0.01 | 0.27 | 0.27 |
| Short-chain saturated fatty acids (SCSFA) (g) | 0.03 | 0.23 | **0.85** | -0.08 | 0.43 | -0.13 | 0.54 | -0.16 |
| Medium-chain saturated fatty acids (MCSFA) (g) | 0.02 | 0.25 | **0.77** | -0.06 | 0.33 | -0.10 | 0.41 | -0.12 |
| Long-chain saturated fatty acids (LCSFA) (g) | 0.17 | **0.79** | 0.50 | -0.04 | **0.86** | 0.04 | 0.32 | -0.06 |
| Long-chain monounsaturated fatty acids (LCMFA) (g) | 0.25 | **0.93** | 0.17 | 0.01 | **0.89** | 0.25 | 0.02 | 0.01 |
| Linoleic Acid (g) | 0.23 | **0.86** | 0.03 | 0.04 | **0.79** | 0.27 | -0.13 | 0.02 |
| Linolenic Acid (g) | 0.25 | **0.70** | 0.16 | 0.06 | **0.64** | 0.28 | -0.07 | 0.02 |
| Arachidonic Acid (g) | 0.24 | 0.47 | -0.05 | 0.40 | 0.51 | 0.02 | 0.09 | 0.54 |
| Eicosapentaenoic Acid (EPA) (g) | 0.06 | 0.01 | 0.07 | **0.88** | 0.00 | 0.09 | 0.04 | **0.84** |
| Docosapentaenoic acid (DPA) (g) | 0.14 | 0.10 | -0.05 | **0.83** | 0.08 | 0.06 | 0.01 | **0.90** |
| Docosahexaenoic Acid (DHA) (g) | 0.10 | 0.10 | 0.03 | **0.98** | 0.02 | 0.06 | 0.05 | **0.90** |
| Total Trans Fatty Acids (g) | 0.05 | **0.60** | 0.34 | -0.03 | **0.62** | -0.04 | 0.16 | -0.06 |
| Total Sugars (g) | 0.33 | 0.25 | 0.37 | -0.02 | 0.40 | 0.22 | 0.37 | -0.08 |
| Starch (g) | 0.52 | 0.51 | 0.13 | -0.04 | **0.64** | 0.49 | 0.08 | -0.07 |
| Calcium (mg) | 0.49 | 0.14 | **0.72** | 0.01 | 0.34 | 0.29 | **0.68** | -0.05 |
| Phosphorus (mg) | **0.68** | 0.43 | 0.44 | 0.17 | 0.59 | 0.49 | 0.48 | 0.16 |
| Magnesium (mg) | **0.83** | 0.27 | 0.23 | 0.10 | 0.38 | **0.76** | 0.30 | 0.09 |

| | | | | | | | | |
|---|---|---|---|---|---|---|---|---|
| Iron (mg) | **0.74** | 0.38 | 0.16 | 0.01 | 0.44 | 0.53 | 0.41 | 0.01 |
| Zinc (mg) | **0.69** | 0.47 | 0.23 | -0.01 | 0.56 | 0.42 | 0.42 | 0.07 |
| Copper (mg) | **0.74** | 0.28 | 0.11 | 0.10 | 0.12 | 0.41 | 0.42 | 0.06 |
| Selenium (mcg) | 0.53 | **0.62** | 0.17 | 0.28 | **0.75** | 0.28 | 0.22 | 0.31 |
| Sodium (mg) | 0.45 | **0.63** | 0.18 | 0.07 | **0.76** | 0.30 | 0.13 | 0.08 |
| Potassium (mg) | **0.78** | 0.27 | 0.27 | 0.12 | 0.40 | **0.72** | 0.33 | 0.15 |
| Manganese (mg) | **0.71** | 0.22 | 0.08 | 0.04 | 0.05 | 0.15 | 0.01 | 0.00 |
| Thiamin (vitamin B1) (mg) | **0.66** | 0.48 | 0.25 | -0.03 | **0.61** | 0.47 | 0.36 | -0.03 |
| Riboflavin (vitamin B2) (mg) | **0.60** | 0.31 | 0.53 | 0.03 | 0.42 | 0.30 | **0.74** | 0.04 |
| Niacin (vitamin B3) (mg) | **0.66** | 0.48 | 0.04 | 0.15 | 0.54 | 0.37 | 0.37 | 0.22 |
| Pantothenic Acid (vitamin B5) (mg) | **0.70** | 0.34 | 0.29 | 0.15 | 0.39 | 0.42 | 0.54 | 0.18 |
| Vitamin B6 (mg) | **0.78** | 0.31 | 0.09 | 0.10 | 0.07 | 0.25 | 0.44 | 0.09 |
| Vitamin B12 (mcg) | 0.47 | 0.20 | 0.40 | 0.21 | -0.09 | 0.05 | 0.41 | 0.06 |
| Natural Folate (mcg) | **0.68** | 0.20 | 0.14 | 0.06 | 0.21 | **0.80** | 0.11 | 0.02 |
| Vitamin C (mg) | 0.45 | 0.03 | 0.08 | 0.07 | 0.02 | 0.44 | 0.15 | 0.06 |
| Retinol (mcg) | 0.31 | 0.09 | **0.62** | 0.06 | -0.01 | 0.09 | **0.62** | 0.02 |
| Beta Carotene Equivalents (mcg) | 0.34 | -0.02 | 0.05 | 0.10 | -0.08 | 0.39 | 0.00 | 0.14 |
| Lutein + Zeaxanthin (mcg) | 0.37 | 0.09 | 0.09 | 0.13 | -0.04 | 0.38 | 0.06 | 0.12 |
| Lycopene (mcg) | 0.17 | 0.15 | 0.06 | -0.01 | 0.21 | 0.26 | 0.05 | -0.05 |
| Vitamin D (mcg) | 0.33 | 0.07 | 0.56 | 0.31 | 0.14 | 0.13 | 0.59 | 0.44 |
| Natural Alpha-Tocopherol (mg) | 0.47 | **0.61** | 0.08 | 0.15 | 0.54 | 0.54 | -0.02 | 0.11 |
| Soluble Dietary Fiber (g) | **0.61** | 0.14 | 0.18 | 0.01 | 0.16 | **0.72** | 0.14 | -0.05 |
| Insoluble Dietary Fiber (g) | **0.75** | 0.15 | 0.08 | 0.02 | 0.20 | **0.86** | 0.08 | -0.02 |
| Proportion of VAR explained (%) | 25.8 | 17.7 | 10.5 | 7.4 | 22.1 | 16.4 | 11.3 | 8.7 |
| Cumulative VAR explained (%) | 25.8 | 43.5 | 50.9 | 61.4 | 22.1 | 38.5 | 49.8 | 58.5 |

[a]Estimated from the frequentist multi-study factor analysis and standard principal component factor analysis carried out on forty-two nutrients. The magnitude of each loading measures the importance of the corresponding nutrient to the factor. [b]Loadings ≥0.6 in absolute value define the dominant nutrients for each factor and were shown in bold typeface. [c]Units of the nutrients indicated their original scale, but loadings

were derived from log-transformed and standardized nutrient intakes entered into the frequentist multi-study factor analysis model and the principal component factor analysis model.

**Supplemental Table 6. Factor-loading matrices and explained variances[a] for the 12 ethnic-background-site-specific dietary patterns identified with the frequentist multi-study factor analysis. Hispanic Community Health Study/Study of Latinos, 2008-2011.**

| Nutrient | BX D | BX CA | CHI CA | MIA CA | MIA Cu | BX M | CHI M | SD M | BX PR | CHI PR | CHI SA | MIA SA |
|---|---|---|---|---|---|---|---|---|---|---|---|---|
| Animal Protein[b,c] (g) | **0.51** | **0.44** | **0.43** | **0.49** | **0.54** | **0.41** | **0.38** | **0.33** | **0.47** | **0.39** | **0.56** | **0.57** |
| Vegetable Protein (g) | -0.25 | -0.18 | **-0.40** | **-0.36** | **-0.37** | **-0.34** | **-0.39** | **-0.35** | **-0.31** | -0.25 | **-0.35** | **-0.40** |
| Cholesterol (mg) | **0.38** | **0.32** | 0.24 | **0.33** | **0.43** | 0.19 | 0.25 | 0.19 | **0.34** | **0.32** | **0.46** | **0.44** |
| SCSFA (g) | 0.06 | -0.08 | 0.13 | -0.06 | 0.11 | 0.01 | 0.14 | 0.04 | 0.02 | 0.07 | 0.05 | 0.03 |
| MCSFA (g) | 0.04 | -0.13 | 0.04 | -0.08 | 0.07 | -0.05 | 0.10 | -0.02 | -0.05 | 0.01 | 0 | -0.02 |
| LCSFA (g) | 0.09 | -0.02 | 0.08 | -0.02 | 0.10 | 0.02 | 0.07 | -0.01 | 0.07 | 0.05 | 0.18 | 0.13 |
| LCMFA (g) | -0.09 | -0.09 | -0.08 | -0.14 | -0.11 | -0.17 | -0.06 | -0.16 | -0.10 | -0.15 | 0.03 | 0.01 |
| Linoleic Acid (g) | -0.13 | -0.11 | -0.19 | -0.18 | -0.19 | -0.28 | -0.14 | -0.23 | -0.21 | -0.26 | -0.04 | -0.11 |
| Linolenic Acid (g) | -0.12 | -0.16 | -0.17 | -0.23 | -0.21 | -0.23 | -0.11 | -0.25 | -0.23 | -0.18 | -0.13 | -0.14 |
| Arachidonic Acid (g) | **0.36** | **0.42** | 0.24 | **0.34** | **0.38** | **0.33** | 0.23 | 0.18 | **0.38** | 0.25 | **0.33** | **0.42** |
| Eicosapentaenoic Acid (EPA) (g) | -0.10 | 0.06 | -0.02 | -0.12 | -0.11 | -0.04 | -0.03 | -0.05 | -0.09 | 0.04 | -0.03 | -0.11 |
| Docosapentaenoic acid (DPA) (g) | 0.12 | 0.28 | 0.10 | 0.09 | 0.07 | 0.14 | 0.05 | 0.02 | 0.12 | 0.07 | 0.01 | 0.1 |
| Docosahexaenoic Acid (DHA) (g) | 0 | 0.18 | 0.04 | -0.01 | -0.01 | 0.08 | 0.03 | -0.01 | 0.03 | 0.09 | 0 | -0.03 |
| Total Trans Fatty Acids (g) | 0.15 | -0.05 | 0.13 | -0.03 | 0.04 | 0.06 | 0.12 | 0.02 | 0.05 | 0.02 | 0.14 | 0.1 |
| Total Sugars (g) | -0.11 | -0.12 | 0.01 | -0.12 | -0.06 | 0 | 0.07 | -0.02 | -0.09 | -0.04 | -0.08 | -0.05 |
| Starch (g) | -0.20 | -0.13 | -0.22 | -0.17 | -0.27 | -0.21 | -0.25 | -0.10 | -0.25 | -0.17 | -0.08 | -0.26 |
| Calcium (mg) | 0.03 | 0.02 | 0 | 0.03 | 0.18 | 0.09 | 0.05 | 0.03 | -0.01 | 0.15 | 0.05 | 0.05 |
| Phosphorus (mg) | 0.12 | 0.15 | -0.04 | 0.06 | 0.17 | 0.01 | -0.13 | 0 | 0.10 | 0.13 | 0.14 | 0.17 |
| Magnesium (mg) | -0.25 | -0.16 | -0.28 | -0.21 | -0.12 | -0.21 | **-0.33** | -0.26 | -0.18 | -0.13 | -0.20 | -0.14 |
| Iron (mg) | 0.06 | 0.11 | 0.01 | 0.07 | -0.08 | 0.05 | 0.08 | 0.19 | 0.01 | 0.03 | 0.02 | -0.05 |
| Zinc (mg) | 0.24 | 0.24 | 0.12 | 0.24 | 0.26 | 0.20 | 0.11 | 0.21 | 0.21 | 0.21 | 0.26 | 0.24 |
| Copper (mg) | -0.20 | -0.05 | -0.23 | -0.07 | -0.12 | -0.14 | -0.21 | -0.22 | -0.22 | -0.16 | -0.14 | -0.19 |
| Selenium (mcg) | 0.25 | 0.23 | 0.14 | 0.19 | 0.20 | 0.24 | 0.15 | 0.10 | 0.18 | 0.17 | 0.24 | 0.22 |

| Nutrient | BX D | BX CA | CHI CA | MIA CA | MIA Cu | BX M | CHI M | SD M | BX PR | CHI PR | CHI SA | MIA SA |
|---|---|---|---|---|---|---|---|---|---|---|---|---|
| Sodium (mg) | 0.06 | -0.02 | 0.05 | 0 | -0.05 | 0.01 | 0.09 | 0.02 | 0.02 | -0.01 | 0.02 | 0.03 |
| Potassium (mg) | -0.29 | -0.19 | -0.14 | -0.18 | -0.03 | -0.15 | -0.16 | -0.23 | -0.16 | -0.13 | -0.16 | 0 |
| Manganese (mg) | -0.24 | -0.18 | **-0.31** | -0.21 | -0.29 | -0.18 | -0.22 | -0.24 | -0.26 | -0.22 | -0.25 | -0.20 |
| Thiamin (vitamin B1) (mg) | 0.02 | 0.03 | -0.02 | 0 | -0.04 | 0.03 | 0.09 | 0.12 | -0.03 | 0.04 | -0.06 | -0.05 |
| Riboflavin (vitamin B2) (mg) | 0.18 | 0.15 | 0.21 | 0.27 | 0.30 | 0.17 | 0.32 | 0.30 | 0.13 | 0.26 | 0.17 | 0.17 |
| Niacin (vitamin B3) (mg) | 0.29 | 0.26 | **0.34** | 0.29 | 0.19 | 0.26 | **0.32** | **0.30** | 0.21 | 0.16 | 0.24 | 0.16 |
| Pantothenic Acid (vitamin B5) (mg) | 0.11 | 0.25 | 0.18 | 0.19 | 0.19 | 0.16 | 0.24 | 0.12 | 0.08 | 0.13 | 0.12 | 0.20 |
| Vitamin B6 (mg) | 0.07 | 0.13 | 0.28 | 0.18 | 0.15 | 0.20 | 0.17 | 0.22 | 0.09 | 0.10 | 0.10 | 0.13 |
| Vitamin B12 (mcg) | 0.29 | 0.23 | **0.40** | **0.43** | **0.42** | **0.40** | **0.46** | **0.44** | 0.27 | **0.4** | **0.43** | 0.24 |
| Natural Folate (mcg) | **-0.42** | **-0.35** | **-0.42** | **-0.38** | **-0.31** | **-0.33** | **-0.37** | **-0.43** | **-0.33** | -0.29 | **-0.41** | -0.21 |
| Vitamin C (mg) | **-0.36** | -0.25 | 0.03 | -0.17 | -0.16 | -0.04 | -0.01 | -0.20 | -0.24 | -0.20 | -0.28 | -0.08 |
| Retinol (mcg) | 0.20 | 0.18 | 0.17 | 0.20 | 0.23 | 0.19 | **0.31** | 0.28 | 0.12 | **0.31** | 0.28 | 0.10 |
| Beta Carotene Equivalents (mcg) | **-0.35** | **-0.37** | -0.01 | -0.18 | -0.12 | -0.13 | -0.02 | -0.27 | -0.26 | -0.19 | -0.20 | -0.03 |
| Lutein + Zeaxanthin (mcg) | -0.26 | -0.29 | -0.06 | -0.12 | -0.05 | -0.16 | 0 | -0.26 | -0.21 | -0.14 | -0.25 | -0.04 |
| Lycopene (mcg) | -0.17 | -0.16 | 0.09 | -0.06 | -0.14 | 0.03 | -0.01 | -0.12 | -0.13 | -0.15 | -0.12 | -0.11 |
| Vitamin D (mcg) | 0.12 | 0.15 | 0.11 | 0.17 | 0.25 | 0.13 | 0.27 | 0.26 | 0.13 | **0.34** | 0.26 | 0.10 |
| Natural Alpha-Tocopherol (mg) | **-0.36** | -0.27 | -0.21 | -0.29 | -0.24 | **-0.33** | -0.20 | **-0.37** | **-0.36** | **-0.32** | -0.28 | -0.18 |
| Soluble Dietary Fiber (g) | **-0.42** | **-0.36** | **-0.39** | **-0.45** | -0.25 | **-0.38** | **-0.48** | **-0.47** | **-0.38** | -0.29 | **-0.44** | -0.28 |
| Insoluble Dietary Fiber (g) | **-0.52** | **-0.47** | **-0.5** | **-0.48** | **-0.41** | **-0.40** | **-0.53** | **-0.45** | **-0.43** | **-0.41** | **-0.53** | **-0.42** |
| Proportion of VAR explained (%) | 14.8 | 16.7 | 10.3 | 14.8 | 11.7 | 11.4 | 8.4 | 15.0 | 12.9 | 9.3 | 12.5 | 10.1 |
| Cumulative VAR explained (%) | 76.2 | 78.1 | 71.7 | 76.2 | 73.1 | 72.8 | 69.8 | 76.4 | 74.3 | 70.7 | 73.9 | 71.5 |

[a]Estimated from the frequentist multi-study factor analysis carried out on 42 nutrients. The magnitude of each loading measures the importance of the corresponding nutrient to the factor. [b]Loadings ≥0.3 in absolute value define the dominant nutrients for each factor and were shown in bold typeface. [c]Units of the nutrients indicated their original scale, but loadings were derived from log-transformed and standardized nutrient intakes entered into the frequentist multi-study factor analysis model.

ABBREVIATIONS: BX: BRONX; CA: Central American; Cu: Cuban; CHI: Chicago; D: Dominican; M: Mexican; MIA: Miami; PR: Puerto Rican; SA: South American; SD: San Diego.

**Supplemental Table 7. Factor congruence coefficients[a] between pairs of ethnic-background-site-specific dietary patterns identified with the frequentist multi-study factor analysis. Hispanic Community Health Study/Study of Latinos, 2008-2011.**

| | BX D | BX CA | CHI CA | MIA CA | MIA Cu | BX M | CHI M | SD M | BX PR | CHI PR | CHI SA | MIA SA |
|---|---|---|---|---|---|---|---|---|---|---|---|---|
| **BX D** | 1 | **0.93** | **0.86** | **0.94** | **0.90** | **0.91** | **0.85** | **0.92** | ***0.97*** | **0.92** | ***0.96*** | **0.88** |
| **BX CA** | | 1 | 0.80 | **0.92** | **0.85** | **0.89** | 0.78 | **0.87** | **0.93** | **0.90** | **0.90** | 0.84 |
| **CHI CA** | | | 1 | **0.92** | **0.89** | ***0.95*** | ***0.96*** | **0.90** | **0.89** | **0.87** | **0.88** | **0.88** |
| **MIA CA** | | | | 1 | **0.93** | ***0.96*** | **0.90** | ***0.95*** | ***0.96*** | **0.94** | **0.94** | **0.90** |
| **MIA Cu** | | | | | 1 | **0.91** | **0.88** | **0.86** | **0.93** | ***0.95*** | **0.93** | ***0.95*** |
| **BX M** | | | | | | 1 | **0.92** | **0.94** | **0.94** | **0.93** | **0.90** | **0.88** |
| **CHI M** | | | | | | | 1 | **0.91** | **0.85** | **0.89** | **0.88** | 0.84 |
| **SD M** | | | | | | | | 1 | **0.92** | **0.94** | **0.90** | 0.79 |
| **BX PR** | | | | | | | | | 1 | **0.93** | **0.94** | **0.92** |
| **CHI PR** | | | | | | | | | | 1 | **0.93** | **0.87** |
| **CHI SA** | | | | | | | | | | | 1 | **0.91** |
| **MIA SA** | | | | | | | | | | | | 1 |

[a]Congruence coefficients range between 0 and 1 (in absolute value), with values between 0.85 and 0.94 indicating fair similarity (in bold typeface in the upper triangular matrix) and values ≥0.95 (in bold and italics typeface in the upper triangular matrix) indicating equivalence of corresponding dietary patterns.

ABBREVIATIONS: BX: BRONX; CA: Central American; Cu: Cuban; CHI: Chicago; D: Dominican; M: Mexican; MIA: Miami; PR: Puerto Rican; SA: South American; SD: San Diego.

**Supplemental Table 8. Factor congruence coefficients[a] between pairs of ethnic-background-site-specific dietary patterns obtained with the Bayesian and frequentist multi-study factor analysis in comparison. Hispanic Community Health Study/Study of Latinos, 2008-2011.**

| | | Frequentist multi-study factor analysis (FMSFA) | | | | | | | | | | | |
|---|---|---|---|---|---|---|---|---|---|---|---|---|---|
| | | **BX D** | **BX CA** | **CHI CA** | **MIA CA** | **MIA Cu** | **BX M** | **CHI M** | **SD M** | **BX PR** | **CHI PR** | **CHI SA** | **MIA SA** |
| **Bayesian multi-study factor analysis (BMSFA)** | **BX D** | ***0.88*** | **0.89** | 0.69 | 0.84 | 0.72 | 0.77 | 0.65 | 0.76 | **0.85** | 0.72 | 0.82 | 0.78 |
| | **BX CA** | 0.71 | ***0.85*** | 0.55 | 0.74 | 0.57 | 0.69 | 0.50 | 0.65 | 0.72 | 0.60 | 0.65 | 0.62 |
| | **CHI CA** | 0.58 | 0.62 | *0.63* | 0.66 | 0.61 | 0.65 | 0.49 | 0.46 | 0.65 | 0.48 | 0.60 | 0.75 |
| | **MIA CA** | 0.73 | 0.80 | 0.70 | *0.83* | 0.69 | 0.76 | 0.66 | 0.71 | 0.75 | 0.65 | 0.74 | 0.75 |
| | **MIA Cu** | 0.62 | 0.70 | 0.55 | 0.69 | *0.64* | 0.62 | 0.51 | 0.53 | 0.63 | 0.55 | 0.65 | 0.73 |
| | **BX M** | 0.64 | 0.73 | 0.74 | 0.80 | 0.68 | *0.79* | 0.68 | 0.67 | 0.71 | 0.63 | 0.66 | 0.74 |
| | **CHI M** | 0.75 | 0.73 | **0.86** | 0.84 | 0.72 | 0.82 | ***0.88*** | 0.80 | 0.73 | 0.71 | 0.77 | 0.76 |
| | **SD M** | 0.80 | 0.81 | 0.77 | **0.86** | 0.68 | 0.83 | 0.78 | ***0.91*** | 0.78 | 0.77 | 0.77 | 0.64 |
| | **BX PR** | 0.76 | 0.81 | 0.67 | 0.80 | 0.71 | 0.74 | 0.58 | 0.64 | *0.82* | 0.64 | 0.74 | 0.82 |
| | **CHI PR** | 0.41 | 0.45 | 0.20 | 0.31 | 0.12 | 0.31 | 0.25 | 0.46 | 0.29 | *0.32* | 0.30 | 0.05 |
| | **CHI SA** | 0.80 | 0.83 | 0.70 | 0.82 | 0.68 | 0.74 | 0.66 | 0.75 | 0.77 | 0.68 | *0.82* | 0.74 |
| | **MIA SA** | 0.63 | 0.65 | 0.59 | 0.65 | 0.61 | 0.60 | 0.50 | 0.46 | 0.66 | 0.48 | 0.62 | *0.78* |

[a]Congruence coefficients range between 0 and 1 (in absolute value), with values between 0.85 and 0.94 indicating “fair similarity” of the corresponding dietary patterns (in bold typeface). The main diagonal (values in italics typeface) provides the comparison between the Bayesian and frequentist version of the same ethnic-background-site-specific dietary pattern.

ABBREVIATIONS: BX: BRONX; CA: Central American; Cu: Cuban; CHI: Chicago; D: Dominican; M: Mexican; MIA: Miami; PR: Puerto Rican; SA: South American; SD: San Diego.

**Supplemental Table 9. Weighted percentages (and standard error) of EBS-specific individuals in the top-quintile category of each shared dietary pattern[a]. Hispanic Community Health Study/Study of Latinos, 2008-2011.**

| | ***Plant-based foods*** | | ***Processed foods*** | | ***Dairy products*** | | ***Seafood*** | |
|---|---|---|---|---|---|---|---|---|
| | **Percentage** | **SE** | **Percentage** | **SE** | **Percentage** | **SE** | **Percentage** | **SE** |
| **BX D** | **18.3** | 1.3 | 22.7 | 1.7 | **22.9** | 1.8 | 19.1 | 1.6 |
| **BX CA** | 22.2 | 4.5 | **18.2** | 3.9 | 22.3 | 4.3 | 20.4 | 4.3 |
| **CHI CA** | 20.9 | 3.3 | 20.3 | 2.5 | 17.9 | 3.3 | 19.5 | 2.8 |
| **MIA CA** | 19.1 | 1.3 | 21.5 | 2.0 | 18.3 | 1.5 | 21.6 | 1.9 |
| **MIA Cu** | 21.4 | 1.1 | 16.5 | 1.0 | 21.5 | 1.3 | 20.0 | 1.0 |
| **BX M** | **24.8** | 4.2 | 18.4 | 4.1 | 22.1 | 4.9 | **13.1** | 3.1 |
| **CHI M** | 20.1 | 1.1 | **23.2** | 1.1 | **17.2** | 0.9 | 16.8 | 0.9 |
| **SD M** | 18.4 | 1.0 | 20.9 | 1.2 | 19.1 | 1.1 | 23.7 | 1.1 |
| **BX PR** | 21.5 | 1.6 | 19.4 | 1.4 | 20.2 | 1.6 | 17.8 | 1.5 |
| **CHI PR** | 20.6 | 1.9 | 19.1 | 2.2 | 18.6 | 1.8 | 16.5 | 2.0 |
| **CHI SA** | 17.4 | 2.6 | 18.6 | 2.7 | 20.1 | 2.8 | **24.3** | 3.6 |
| **MIA SA** | 18.5 | 2.6 | 21.2 | 2.6 | 18.1 | 2.2 | 18.7 | 2.1 |

[a]For each dietary pattern, minimum and maximum percentages are indicated in bold typeface.
ABBREVIATIONS: BX: BRONX; CA: Central American; Cu: Cuban; CHI: Chicago; D: Dominican; M: Mexican; MIA: Miami; PR: Puerto Rican; SA: South American; SD: San Diego; SE: standard error.

**Supplemental Table 10. Socio-demographic and lifestyle characteristics (weighted proportion and standard error in parenthesis) by ethnic-background-site-specific tertile-based (Q1, Q3) categories. Hispanic Community Health Study/Study of Latinos, 2008-2011.**

| | BX D | | | BX CA | | | CHI CA | | | MIA CA | | | MIA Cub | | |
|---|---|---|---|---|---|---|---|---|---|---|---|---|---|---|---|
| | Q1 | Q3 | p-value[a] | Q1 | Q3 | p-value[a] | Q1 | Q3 | p-value[a] | Q1 | Q3 | p-value[a] | Q1 | Q3 | p-value[a] |
| **Age (y) (%)** | | | | | | | | | | | | | | | |
| 18-44 | 0.33 (0.05) | 0.47 (0.04) | | 0.35 (0.12) | 0.6 (0.09) | | 0.38 (0.08) | 0.62 (0.06) | | 0.51 (0.05) | 0.50 (0.06) | | 0.30 (0.03) | 0.35 (0.02) | |
| 45-55 | 0.33 (0.04) | 0.25 (0.03) | <.001 | 0.45 (0.14) | 0.23 (0.07) | 0.96 | 0.25 (0.06) | 0.22 (0.05) | 0.72 | 0.24 (0.03) | 0.29 (0.05) | 0.09 | 0.27 (0.02) | 0.28 (0.02) | <.001 |
| 55-74 | 0.34 (0.04) | 0.28 (0.04) | | 0.20 (0.07) | 0.16 (0.06) | | 0.37 (0.07) | 0.16 (0.04) | | 0.26 (0.04) | 0.21 (0.04) | | 0.44 (0.02) | 0.38 (0.03) | |
| **Sex (%)** | | | | | | | | | | | | | | | |
| Female | 0.66 (0.04) | 0.50 (0.04) | 0.01 | 0.62 (0.15) | 0.41 (0.10) | 0.96 | 0.53 (0.06) | 0.33 (0.05) | 0.52 | 0.60 (0.04) | 0.45 (0.04) | 0.01 | 0.53 (0.03) | 0.38 (0.02) | <.001 |
| **Immigrant generation (%)** | | | | | | | | | | | | | | | |
| First | 0.88 (0.02) | 0.78 (0.03) | 0.17 | 0.88 (0.05) | 0.75 (0.08) | 0.96 | 0.94 (0.03) | 0.92 (0.04) | 0.72 | 0.98 (0.01) | 0.95 ().02) | 0.56 | 0.94 (0.02) | 0.93 (0.01) | 0.79 |
| **Years lived in mainland US (50 states and DC) (%)** | | | | | | | | | | | | | | | |
| Born in mainland United States | 0.24 (0.03) | 0.24 (0.03) | | 0.26 (0.08) | 0.12 (0.04) | | 0.32 (0.07) | 0.38 (0.06) | | 0.46 (0.03) | 0.38 (0.04) | | 0.53 (0.02) | 0.48 (0.03) | |
| < 10 ys in US | 0.64 (0.03) | 0.54 (0.03) | 0.25 | 0.62 (0.09) | 0.62 (0.08) | 0.96 | 0.62 (0.06) | 0.55 (0.06) | 0.72 | 0.53 (0.03) | 0.57 (0.04) | 0.41 | 0.41 (0.02) | 0.45 (0.02) | 0.79 |
| ≥10 ys in US | 0.12 (0.03) | 0.22 (0.03) | | 0.12 (0.05) | 0.25 (0.08) | | 0.06 (0.03) | 0.08 (0.04) | | 0.01 (0.01) | 0.05 (0.02) | | 0.06 (0.02) | 0.07 (0.01) | |
| **Age of immigration[b] (y)** | 31.8 (0.79) | 28.9 (1.06) | 0.04 | 26.8 (1.51) | 24.4 (1.94) | 0.99 | 30.5 (1.34) | 25.7 (1.26) | 0.72 | 33.2 (1.0) | 29.5 (1.17) | 0.01 | 39.1 (0.8) | 38.2 (1.01) | 0.79 |
| **Employment (%)** | | | | | | | | | | | | | | | |

| | | | | | | | | | | | | | | | |
|---|---|---|---|---|---|---|---|---|---|---|---|---|---|---|---|
| Retired and not currently employed | 0.16 (0.03) | 0.11 (0.02) | | 0.12 (0.05) | 0.06 (0.03) | | 0.12 (0.06) | 0.04 (0.02) | | 0.04 (0.02) | 0.03 (0.02) | | 0.16 (0.02) | 0.13 (0.02) | |
| Not retired and not currently employed | 0.32 (0.04) | 0.35 (0.03) | 0.25 | 0.21 (0.07) | 0.30 (0.09) | 0.99 | 0.30 (0.05) | 0.27 (0.06) | 0.72 | 0.31 (0.04) | 0.40 (0.05) | 0.23 | 0.43 (0.03) | 0.46 (0.03) | 0.79 |
| Part-time (≤35 hrs) | 0.14 (0.02) | 0.18 (0.03) | | 0.39 (0.13) | 0.26 (0.10) | | 0.25 (0.06) | 0.19 (0.05) | | 0.23 (0.04) | 0.23 (0.03) | | 0.11 (0.02) | 0.09 (0.02) | |
| Full-time (35+hrs) | 0.38 (0.05) | 0.36 (0.03 | | 0.28 (0.15) | 0.38 (0.10) | | 0.33 (0.06) | 0.50 (0.07) | | 0.42 (0.03) | 0.34 (0.05) | | 0.30 (0.02) | 0.32 (0.03) | |
| **Marital Status (%)** | | | | | | | | | | | | | | | |
| Single | 0.20 (0.03) | 0.37 (0.04) | | 0.31 (0.11) | 0.27 (0.10) | | 0.14 (0.06) | 0.15 (0.04) | | 0.26 (0.04) | 0.27 (0.03) | | 0.15 (0.02) | 0.20 (0.02) | |
| Married/living with partner | 0.53 (0.04) | 0.44 (0.04) | 0.01 | 0.51 (().13) | 0.55 (0.10) | 0.96 | 0.72 (0.07) | 0.71 (0.05) | 0.72 | 0.54 (0.04) | 0.49 (0.04) | 0.61 | 0.57 (0.03) | 0.59 (0.02) | 0.79 |
| Separated/divorced/widowed | 0.26 (0.04) | 0.20 (0.03) | | 0.18 (0.07) | 0.18 (0.06) | | 0.14 (0.04) | 0.13 (0.03 | | 0.20 (0.03) | 0.24 (0.03) | | 0.28 (0.02) | 0.22 (0.02) | |
| **Yearly household income (%)** | | | | | | | | | | | | | | | |
| < $30k | 0.68 (0.04) | 0.78 (0.03) | | 0.55 (0.10) | 0.54 (0.11) | | 0.72 (0.06) | 0.68 (0.07) | | 0.71 (0.03) | 0.74 (0.04) | | 0.6 7 (0.04) | 0.68 (0.03) | |
| ≥ $30k | 0.28 (0.03) | 0.18 (0.03) | 0.31 | 0.42 (0.11) | 0.34 (0.11) | 0.96 | 0.27 (0.06) | 0.31 (0.07) | 0.72 | 0.20 (0.04) | 0.21 (0.04) | 0.84 | 0.22 (0.02) | 0.21 (0.03) | 0.80 |
| Missing | 0.04 (0.01) | 0.04 (0.01) | | 0.03 (0.03) | 0.12 (0.07) | | 0.01 (0.01) | 0.01 (0.01) | | 0.09 (0.02) | 0.05 (0.02) | | 0.10 (0.02) | 0.10 (0.02) | |
| **Education status (%)** | | | | | | | | | | | | | | | |
| Less than high school education | 0.43 (0.04) | 0.52 (0.04) | | 0.35 (0.11) | 0.42 (0.10) | | 0.46 (0.07) | 0.64 (0.07) | | 0.37 (0.04) | 0.37 (0.06) | | 0.19 (0.02) | 0.26 (0.02) | |
| High school education or equivalent | 0.21 (0.03) | 0.17 (0.03) | 0.53 | 0.17 (0.07) | 0.26 (0.09) | 0.99 | 0.13 (0.03) | 0.06 (0.02) | 0.72 | 0.24 (0.03) | 0.29 (0.04) | 0.46 | 0.31 (0.03) | 0.30 (0.03) | 0.34 |
| Greater than high school education | 0.37 (0.04) | 0.31 (0.04) | | 0.49 (0.12) | 0.33 (0.10) | | 0.41 (0.07) | 0.29 (0.06) | | 0.39 (0.04) | 0.34 (0.06) | | 0.51 (0.03) | 0.43 (0.03) | |
| **Met 2008 Physical Activity Guidelines for Americans (%)** | | | | | | | | | | | | | | | |

| | | | | | | | | | | | | | | | |
|---|---|---|---|---|---|---|---|---|---|---|---|---|---|---|---|
| No | 0.48 (0.04) | 0.45 (0.04) | 0.53 | 0.37 (0.10) | 0.47 (0.10) | 0.96 | 0.60 (0.06) | 0.57 (0.07) | 0.72 | 0.70 (0.03) | 0.65 (0.04) | 0.82 | 0.80 (0.02) | 0.78 (0.02) | 0.76 |
| **Supplemental Use (%)** | | | | | | | | | | | | | | | |
| No | 0.53 (0.04) | 0.66 (0.04) | 0.04 | 0.75 (0.08) | 0.50 (0.11) | 0.96 | 0.59 (0.07) | 0.59 (0.06) | 0.72 | 0.49 (0.04) | 0.55 (0.05) | 0.31 | 0.48 (0.03) | 0.60 (0.03) | 0.04 |
| **Body Mass Index[c] (%)** | | | | | | | | | | | | | | | |
| Underweight | 0.01 (0.01) | 0.02 (0.01) | | 0.04 (0.04) | 0.00 (0.00) | | 0.00 (0.0) | 0.00 (0.0) | | 0.02 (0.01) | 0.01 (0.01) | | 0.01 (0.00) | 0.01 (0.00) | |
| Normal | 0.20 (0.03) | 0.20 (0.02) | 0.70 | 0.23 (0.09) | 0.15 (0.05) | 0.99 | 0.19 (0.05 | 0.21 (0.05) | 0.72 | 0.23 (0.03) | 0.21 (0.04) | 0.55 | 0.24 (0.02) | 0.23 (0.02) | 0.79 |
| Overweight | 0.38 (0.03) | 0.35 (0.03) | | 0.35 (0.08) | 0.41 (0.07) | | 0.41 (0.06) | 0.52 (0.06) | | 0.38 (0.03) | 0.37 (0.04) | | 0.39 ().02) | 0.38 (0.02) | |
| Obesity | 0.42 (0.03) | 0.41 (0.03) | | 0.38 (0.09) | 0.44 (0.08) | | 0.30 (0.05) | 0.27 (0.05) | | 0.37 (0.03) | 0.41 (0.03) | | 0.36 (0.02) | 0.38 (0.02) | |
| **Energy[b] (kcal)** | 1501.2 (53.8) | 1444.9 (50.1) | 0.70 | 1584.7 (133.4) | 1621.8 (215.0) | 0.99 | 1871.6 (74.9) | 1905.8 (87.5) | 0.72 | 1869.9 (75.8) | 1932.5 (62.5) | 0.69 | 2056.2 (44.2) | 2102.5 (43.8) | 0.34 |

**Supplemental Table 10. Socio-demographic and lifestyle characteristics by ethnic-background-site-specific tertile-based (Q1, Q3) categories. Hispanic Community Health Study/Study of Latinos, 2008-2011. (continued).**

| | BX M | | | CHI M | | | SD M | | | BX PR | | | CHI PR | | | CHI SA | | | MIA SA | | |
|---|---|---|---|---|---|---|---|---|---|---|---|---|---|---|---|---|---|---|---|---|---|
| | Q1 | Q3 | p-value[a] | Q1 | Q3 | p-value[a] | Q1 | Q3 | p-value[a] | Q1 | Q3 | p-value[a] | Q1 | Q3 | p-value[a] | Q1 | Q3 | p-value[a] | Q1 | Q3 | p-value[a] |
| **Age (y) (%)** | | | | | | | | | | | | | | | | | | | | | |
| 18-44 | 0.71 (0.08) | 0.94 (0.03) | | 0.50 (0.03) | 0.69 (0.03) | | 0.41 (0.04) | 0.47 (0.04) | | 0.25 (0.04) | 0.17 (0.04) | | 0.28 (0.08) | 0.19 (0.05) | | 0.55 (0.08) | 0.50 (0.08) | | 0.36 (0.06) | 0.50 (0.07) | |
| 45-55 | 0.14 (0.06) | 0.03 (0.02) | 0.15 | 0.28 (0.02) | 0.17 (0.02) | <.001 | 0.28 (0.02) | 0.27 (0.03) | <.001 | 0.21 (0.03) | 0.32 (0.08) | 0.40 | 0.14 (0.03) | 0.42 (0.07) | 0.75 | 0.16 (0.04) | 0.25 (0.05) | 0.54 | 0.26 (0.04) | 0.30 (0.05) | 0.43 |
| 55-74 | 0.15 (0.07) | 0.02 (0.02) | | 0.22 (0.02) | 0.14 (0.02) | | 0.31 (0.03) | 0.26 (0.04) | | 0.54 (0.04) | 0.50 (0.07) | | 0.58 (0.08) | 0.40 (0.05) | | 0.29 (0.06) | 0.25 (0.06) | | 0.38 (0.06) | 0.20 (0.05) | |
| **Sex (%)** | | | | | | | | | | | | | | | | | | | | | |
| Female | 0.69 (0.07) | 0.25 (0.07) | 0.01 | 0.49 (0.02) | 0.51 (0.03) | 0.07 | 0.67 ().04) | 0.56 (0.03) | <.001 | 0.58 (0.04) | 0.52 (0.06) | 0.17 | 0.46 (0.07) | 0.54 (0.06) | <.001 | 0.59 (0.07) | 0.28 (0.06) | 0.01 | 0.70 (0.06) | 0.57 (0.07) | 0.43 |
| **Immigrant Generation (%)** | | | | | | | | | | | | | | | | | | | | | |
| First | 0.97 (0.01) | 0.96 (0.03) | 0.49 | 0.90 (0.01) | 0.79 (0.03) | <.001 | 0.73 (0.02) | 0.59 (0.02) | <.001 | 0.53 (0.03) | 0.49 (0.04) | 0.40 | 0.47 (0.04) | 0.45 (0.04) | 0.95 | 0.92 (0.05) | 0.94 (0.03) | 0.17 | 0.97 (0.02) | 0.96 (0.02) | 0.89 |
| **Years lived in mainland US (50 states + DC) (%)** | | | | | | | | | | | | | | | | | | | | | |
| Born in mainland United States | 0.38 (0.08) | 0.37 (0.09) | | 0.22 (0.02) | 0.27 (0.02) | | 0.23 (0.03) | 0.20 (0.02) | | 0.08 (0.02) | 0.06 (0.02) | | 0.05 (0.02) | 0.04 (0.02) | | 0.30 (0.06) | 0.42 (0.06) | | 0.43 (0.06) | 0.47 (0.05) | |
| < 10 ys in US | 0.60 (0.08) | 0.59 (0.09) | 0.64 | 0.69 (0.02) | 0.52 (0.02) | <.001 | 0.55 (0.03) | 0.42 (0.02) | <.001 | 0.47 (0.03) | 0.48 (0.04) | 0.40 | 0.43 (0.04) | 0.43 (0.04) | 0.95 | 0.62 (0.05) | 0.52 (0.06) | 0.20 | 0.54 (0.06) | 0.49 (0.05) | 0.89 |
| ≥ 10 ys in US | 0.03 (0.01) | 0.04 (0.03) | | 0.09 (0.01) | 0.21 (0.03) | | 0.22 (0.02) | 0.39 ().02) | | 0.45 (0.03) | 0.46 (0.03) | | 0.52 (0.04) | 0.53 (0.04) | | 0.08 (0.05) | 0.06 (0.03) | | 0.03 (0.02) | 0.04 (0.02) | |
| **Age of Immigration[b]** | 26.9 (1.3) | 21.8 (1.3) | 0.15 | 26.6 (0.55) | 24.4 (0.5) | 0.07 | 29.0 (1.1) | 29.3 (1.1) | 0.20 | 18.7 (1.2) | 16.9 (2.0) | 0.13 | 21.3 (1.5) | 17.4 (1.5) | 0.75 | 29.8 (1.6) | 30.9 (1.3) | 0.78 | 36.3 (1.4) | 32.4 (1.5) | 0.43 |
| **Employment (%)** | | | | | | | | | | | | | | | | | | | | | |

| | | | | | | | | | | | | | | | | | | | | | |
|---|---|---|---|---|---|---|---|---|---|---|---|---|---|---|---|---|---|---|---|---|---|
| Retired and not currently employed | 0.02 (0.01) | 0.02 (0.02) | | 0.05 (0.01) | 0.04 (0.01) | | 0.07 (0.01) | 0.06 (0.01) | | 0.31 (0.04) | 0.31 (0.05) | | 0.36 (0.08) | 0.21 (0.04) | | 0.04 (0.02) | 0.04 (0.02) | | 0.10 (0.05) | 0.04 (0.02) | |
| Not retired and not currently employed | 0.43 (0.08) | 0.18 (0.07) | 0.15 | 0.34 (0.02) | 0.29 (0.03) | 0.04 | 0.32 (0.03) | 0.40 (0.04) | 0.20 | 0.38 (0.04) | 0.45 (0.07) | 0.53 | 0.21 (0.05) | 0.43 (0.07) | 0.75 | 0.21 (0.05) | 0.20 (0.06) | 0.93 | 0.29 (0.05) | 0.25 (0.06) | 0.43 |
| Part-time (≤35 hrs) | 0.29 (0.08) | 0.18 (0.08) | | 0.14 (0.02) | 0.20 (0.02) | | 0.21 (0.03) | 0.19 (0.03) | | 0.06 (0.01) | 0.07 (0.02) | | 0.15 (0.06) | 0.08 (0.03) | | 0.32 (0.05) | 0.25 (0.05) | | 0.20 (0.04) | 0.18 (0.05) | |
| Full-time (35+hrs) | 0.27 (0.08) | 0.62 (0.11) | | 0.47 (0.03) | 0.47 (0.03) | | 0.40 (0.04) | 0.36 (0.03) | | 0.25 (0.04) | 0.18 (0.04) | | 0.28 (0.07) | 0.29 (0.06) | | 0.43 (0.08) | 0.52 (0.08) | | 0.41 (0.06) | 0.54 (0.07) | |
| **Marital Status (%)** | | | | | | | | | | | | | | | | | | | | | |
| Single | 0.12 (0.06) | 0.09 (0.05) | | 0.09 (0.02) | 0.13 (0.02) | | 0.12 (0.02) | 0.10 (0.02) | | 0.32 (0.04) | 0.38 (0.08) | | 0.14 (0.04) | 0.24 (0.07) | | 0.16 (0.06) | 0.05 (0.02) | | 0.17 (0.04) | 0.19 (0.05) | |
| Married/living with partner | 0.71 (0.09) | 0.87 (0.06) | 0.50 | 0.76 (0.02) | 0.72 (0.03) | 0.05 | 0.69 (0.03) | 0.72 (0.03) | <.001 | 0.35 (0.04) | 0.36 (0.05) | 0.18 | 0.52 (0.07) | 0.47 (0.06) | 0.75 | 0.69 (0.07) | 0.76 (0.08) | 0.93 | 0.58 (0.06) | 0.58 (0.06) | 0.89 |
| Separated/divorced/widowed | 0.17 (0.07) | 0.04 (0.02) | | 0.15 (0.02) | 0.15 (0.02) | | 0.19 (0.02) | 0.18 (0.02) | | 0.33 (0.04) | 0.26 (0.04) | | 0.34 (0.08) | 0.29 (0.05) | | 0.16 (0.04) | 0.19 (0.08) | | 0.25 (0.05) | 0.24 (0.05) | |
| **Yearly Household Income (%)** | | | | | | | | | | | | | | | | | | | | | |
| < $30k | 0.65 (0.07) | 0.91 (0.04) | | 0.73 (0.02) | 0.62 (0.04) | | 0.52 (0.04) | 0.66 (0.04) | | 0.69 (0.04) | 0.78 (0.04) | | 0.72 (0.07) | 0.60 (0.06) | | 0.56 (0.08) | 0.70 (0.07) | | 0.66 (0.06) | 0.56 (0.06) | |
| ≥$30k | 0.26 (0.06) | 0.06 (0.03) | 0.49 | 0.24 (0.02) | 0.35 (0.03) | 0.05 | 0.47 (0.04) | 0.31 (0.03) | 0.04 | 0.27 (0.04) | 0.19 (0.04) | 0.82 | 0.28 (0.07) | 0.34 (0.05) | 0.75 | 0.40 (0.08) | 0.29 (0.07) | 0.93 | 0.28 (0.06) | 0.42 (0.07) | 0.43 |
| Missing | 0.09 (0.05) | 0.03 (0.02) | | 0.03 (0.01) | 0.03 (0.01) | | 0.02 (0.00) | 0.03 (0.01) | | 0.04 (0.01) | 0.03 (0.01) | | 0.01 (0.01) | 0.05 (0.04) | | 0.04 (0.04) | 0.01 (0.01) | | 0.07 (0.03) | 0.03 (0.02) | |
| **Education status (%)** | | | | | | | | | | | | | | | | | | | | | |
| Less than high school education | 0.59 (0.08) | 0.64 (0.11) | 0.94 | 0.63 (0.03) | 0.51 (0.03) | 0.00 | 0.32 (0.03) | 0.42 (0.04) | 0.02 | 0.46 (0.04) | 0.49 (0.06) | 0.40 | 0.54 (0.07) | 0.47 (0.06) | 0.82 | 0.29 (0.06) | 0.41 (0.08) | 0.01 | 0.15 (0.05) | 0.21 (0.06) | 0.89 |
| High school education or equivalent | 0.33 (0.08) | 0.25 (0.08) | | 0.73 (0.02) | 0.31 (0.03) | | 0.25 (0.03) | 0.24 (0.03) | | 0.22 (0.03) | 0.24 (0.04) | | 0.18 (0.04) | 0.22 (0.05) | | 0.21 (0.06) | 0.27 (0.07) | | 0.28 (0.06) | 0.18 (0.04) | |

| | | | | | | | | | | | | | | | | | | | | | |
|---|---|---|---|---|---|---|---|---|---|---|---|---|---|---|---|---|---|---|---|---|---|
| Greater than high school education | 0.08 (0.05) | 0.11 (0.10) | | 0.14 (0.02) | 0.18 (0.03) | | 0.43 (0.04) | 0.34 (0.03) | | 0.31 (0.04) | 0.27 (0.08) | | 0.30 (0.07) | 0.31 (0.05) | | 0.50 (0.06) | 0.31 (0.06) | | 0.57 (0.06) | 0.61 (0.06) | |
| **Met 2008 Physical Activity Guidelines for Americans (%)** | | | | | | | | | | | | | | | | | | | | | |
| No | 0.25 (0.07) | 0.23 (0.10) | 0.38 | 0.57 (0.02 | 0.61 (0.03) | 0.88 | 0.59 (0.04) | 0.62 (0.04) | 0.26 | 0.55 (0.04) | 0.47 (0.06) | 0.82 | 0.64 (0.07) | 0.72 (0.05) | 0.20 | 0.56 (0.06) | 0.60 (0.07) | 0.95 | 0.70 (0.05) | 0.73 (0.07) | 0.89 |
| **Supplemental Use (%)** | | | | | | | | | | | | | | | | | | | | | |
| No | 0.51 (0.08) | 0.72 (0.11) | 0.49 | 0.67 (0.03) | 0.60 (0.04) | 0.88 | 0.42 (0.04) | 0.47 (0.03) | 0.20 | 0.54 (0.04) | 0.62 (0.05) | 0.17 | 0.50 (0.07) | 0.47 (0.05) | 0.55 | 0.49 (0.07) | 0.51 (0.07) | 0.52 | 0.25 (0.05) | 0.36 (0.07) | <.001 |
| **Body Mass Index[c] (%)** | | | | | | | | | | | | | | | | | | | | | |
| Underweight | 0.00 (0.00) | 0.00 (0.00) | | 0.01 (0.01) | 0.01 (0.00) | | 0.02 (0.01) | 0.00 (0.00) | | 0.01 (0.01) | 0.00 (0.00) | | 0.01 (0.01) | 0.00 (0.00) | | 0.07 (0.05) | 0.00 (0.001) | | 0.00 (0.00) | 0.02 (0.01) | |
| Normal | 0.21 (0.05) | 0.20 (0.06) | NA | 0.22 (0.02) | 0.21 (0.02) | 0.50 | 0.23 (0.02) | 0.20 (0.02) | 0.39 | 0.18 (0.02) | 0.17 (0.02) | 0.79 | 0.25 (0.04) | 0.20 (0.03) | 0.55 | 0.28 (0.05) | 0.22 (0.05) | 0.29 | 0.33 (0.06) | 0.35 (0.04) | 0.71 |
| Overweight | 0.41 (0.06) | 0.29 (0.08) | | 0.39 (0.02) | 0.34 (0.03) | | 0.40 (0.03) | 0.38 (0.03) | | 0.33 (0.03) | 0.32 (0.03) | | 0.25 (0.04) | 0.39 (0.05) | | 0.36 (0.05) | 0.50 (0.06) | | 0.37 (0.06) | 0.37 (0.04) | |
| Obesity | 0.37 (0.07) | 0.51 (0.09) | | 0.38 (0.02) | 0.45 (0.02) | | 0.36 (0.02) | 0.41 (0.03) | | 0.47 (0.03) | 0.51 (0.04) | | 0.49 (0.04) | 0.41 (0.04) | | 0.28 (0.06) | 0.27 (0.06) | | 0.30 (0.05) | 0.26 (0.05) | |
| **Energy (kcal)[a]** | 1691 (116) | 1678 (126) | 0.49 | 1861 (31) | 1859 (39) | 0.03 | 1869 (40) | 1803 (56) | 0.47 | 1505 (65) | 1453 (86) | 0.18 | 1877 (117) | 1930 (103) | 0.55 | 1876 (103) | 1890 (115) | 0.92 | 1881 (64) | 2082 (78) | 0.89 |

[a]P-values are from Chi-square test of independence for categorical characteristics and from ANOVA for continuous characteristics. P-values were adjusted for multiple comparisons using the False Discovery Rate method. The adjustment was carried out separately by each ethnic-background-site-specific dietary pattern.
[b]Values are weighted means with standard errors indicated in parenthesis.
[c]Weight categories were as follows: Underweight (<18.5 kg/m$^2$), Normal (18.5–24.9 kg/m$^2$), Overweight (25–29.9 kg/m$^2$), and Obesity (≥30 kg/m$^2$).

ABBREVIATIONS: AHEI: Alternate Healthy Eating Index; BX: BRONX; CA: Central American; Cu: Cuban; CHI: Chicago; D: Dominican; M: Mexican; MIA: Miami; NA: Not Available; PR: Puerto Rican; SA: South American; SD: San Diego.

**Supplemental Figure 1. Flowchart of baseline study participants overall and by ethnic-background-site-specific category. Hispanic Community Health Study/Study of Latinos, 2008-2011.**

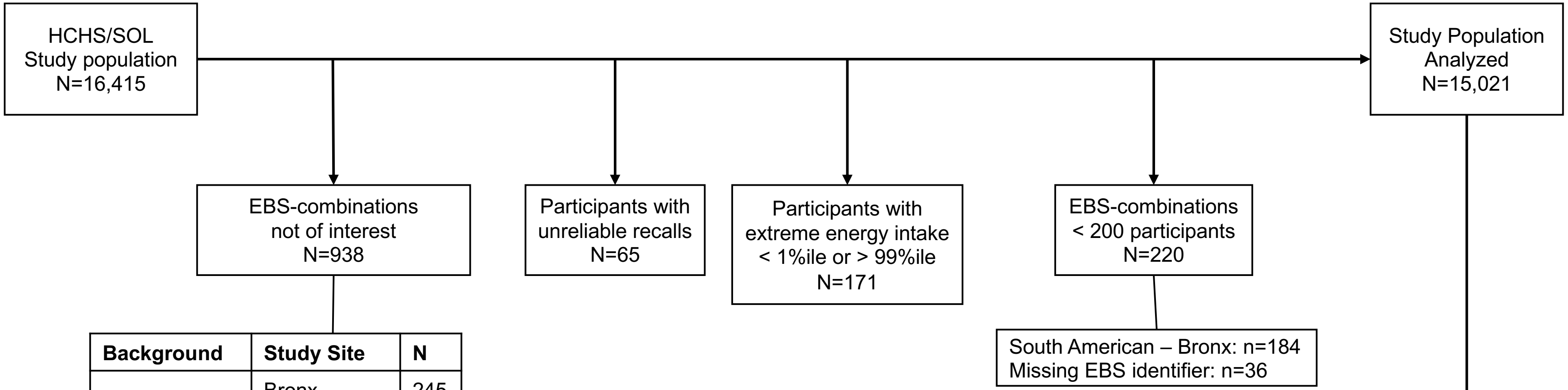


| Background | Study Site | N |
|---|---|---|
| Mixed/Other | Bronx | 245 |
| | Chicago | 152 |
| | Miami | 296 |
| | San Diego | 245 |

| Background | Study Site | Num Recalls 1 | Num Recalls 2 | Total |
|---|---|---|---|---|
| Dominican | Bronx | 228 | 1134 | **1362** |
| Central American | Bronx | 40 | 177 | **217** |
| | Chicago | 13 | 404 | **417** |
| | Miami | 135 | 877 | **1012** |
| Cuban | Miami | 256 | 1985 | **2241** |
| Mexican | Bronx | 25 | 180 | **205** |
| | Chicago | 196 | 2207 | **2403** |
| | San Diego | 469 | 3306 | **3775** |
| Puerto Rican | Bronx | 336 | 1454 | **1790** |
| | Chicago | 71 | 690 | **761** |
| South American | Chicago | 30 | 343 | **373** |
| | Miami | 24 | 441 | **465** |
| | **Total** | **1823** | **13198** | **15021** |

ABBREVIATIONS: EBS: ethnic background site.
